# Intuitive, multidimensional motor unit control after paralysing spinal cord injury using non-invasive recordings

**DOI:** 10.64898/2025.12.09.25341760

**Authors:** Xingchen Yang, Vishal Rawji, Ciara Gibbs, Tianyu Jia, Lara Gouveia Vila, Agnese Grison, Ajoy Nair, Dario Farina, Juan Álvaro Gallego

## Abstract

Spinal cord injury (SCI) results in profound motor impairment for approximately 20 million people worldwide. Regaining hand use is one of their highest priorities. Interestingly, even severely affected individuals can still generate some level of muscle activity (EMG) in their paralysed muscles when attempting to perform a movement. Here we asked whether participants with no hand function due to a cervical SCI could still use their spared inputs to volitionally control the activity of motor units from paralysed muscles, which we detected using high-density surface EMG recordings. Participants could readily control those motor units to accurately perform three different one-dimensional tracking tasks that imposed different control requirements, with their performance plateauing after a few days of practice. Moreover, this accurate control readily transferred to a two-dimensional cursor navigation task and allowed participants to play a series of one-dimensional (“Pong”), and two-dimensional (“Snake”) games, as well as navigate a virtual wheelchair using three independent control signals. This robust performance was achieved despite the presence of spasticity, and even in one participant who had a motor complete injury. These findings demonstrate the promise of non-invasive motor unit recordings for restoring hand function, as well as a potential tool for neurorehabilitation after paralysis.

## Introduction

Spinal cord injury (SCI) affects approximately 20 million people worldwide, leading to major disability^1^. Loss of hand function profoundly reduces independence and quality of life; consequently, tetraplegic individuals with SCI highly prioritise its recovery^2^.

Advances in invasive techniques to stimulate the spinal cord^3^, nerves^4^, or muscles^5–7^ show promising results for restoration of volitional control of the hand by potentiating residual connections, while intracortical brain-computer interfaces^8–10^ (BCIs) hold potential for functional compensation by mapping cortical activity onto signals to control external devices. Yet, in both cases, the need for surgery limits their availability^11^. Here, we explore a non-invasive alternative for functional compensation for lost hand function.

Even following a complete lesion that prevents the execution of any overt movement, there remain residual connections between motor regions in the brain and spinal motoneurons below the injury^12^. As a result, it is possible to detect residual muscle activity (electromyography, EMG) using surface electrodes^13,14^. Previous work provides evidence that people with an incomplete SCI can use these residual EMG signals to control computer cursors^15^ or assistive devices^16,17^. Yet the surface EMG signals from paralysed muscles have inherent limitations that prevent accurate control. First, since the injury deprives many motor units^*^ from their inputs, surface EMG signals in people with SCI have low signal-to-noise ratio, reflective of this small number of active units. This makes them unreliable as control signals^18^. Second, for the up to 70% of SCI survivors suffering from spasticity^19,20^, this signal becomes unusable due to the tonic firing of many of the remaining motor units^21,22^. This would prevent many people from benefitting from this type of technology.

Paradoxically, control based on the activity of a few single motor units —as opposed to the “global” EMG—could overcome these limitations: motor unit action potentials are discrete events that can be counted —like in intracortical BCIs^8,9^— to extract intuitive control signals, and, in people with spasticity, one could potentially filter out motor units that exhibit tonic firing and leave those that can be voluntarily controlled. This is the hypothesis that we address here building on observations that single motor unit activity can be detected from surface EMG recordings in people with SCI^13,14^, enabling tracking of simple target profiles^13,23^, and offline classification of attempted single finger movements^24^.

We investigated the control performance, potential for improvement, and dimensionality of motor unit control in three people with no functional use of their hand due to SCI. Even if they exhibited, at most, minimal flickers of muscle contraction during attempted movements, all participants could readily control the activity of motor units from affected muscles to accurately track three different one-dimensional profiles, even in the presence of significant spasticity. This control improved during the first days of practice, with performance plateauing before five sessions. Remarkably, participants could readily extend this control to a second and even a third degree of freedom, with minimal degradation in control performance. This robust control allowed them to play adapted versions of the games “Pong” (one degree of freedom) and “Snake” (two degrees of freedom), and, in a step towards functional applications, navigate a virtual wheelchair using three degrees of freedom. Our work thus paves the way for future assistive devices and gamified therapies for people with paralysis.

## Results

### Experimental paradigm

Three men (in their 30’s, 60’s and 50’s named P1, P2, and P3) who suffered from a SCI ranging from C1 to C4 level participated in this study. None had any functional movements in their tested hands, with MRC power ranging from 0 (no movement) to 1 (flicker of movement). Detailed participant characteristics are summarised in Table 1 (Methods).

**Table 1.** Participant characteristics.

| Participant | Age | Gender | Level | ASIA<sup>1</sup> | Tested hand | Upper limb MRC power of tested arm<sup>2</sup> | Duration of injury (months) | Experiment time (months) |
| --- | --- | --- | --- | --- | --- | --- | --- | --- |
| P1 | 30's | male | C4 | C | L | 0-1 in hand and wrist* | 24 | 9 |
| P2 | 60's | male | C1-C3 | C | R | 0 in hand and wrist** | 13 | 3 |
| P3 | 50's | male | C2-C4 | A | R | 0-1 in hand and wrist | 12 | 6 |

<sup>1</sup>Scale to measure severity of spinal cord injury. Includes measurement of sensory and motor function. ASIA A = complete injury, ASIA B = sensory but not motor function. C = motor incomplete.
<sup>2</sup>MRC grading scale tests power. Results here report MRC graded power in wrist and hands. 0 = no movement; 1 = flicker of contraction; 2 = active movement with gravity eliminated; 3 = active movement against gravity; 4 = active movement against resistance; 5 = normal power.
\*Except wrist extension 3/5
\*\*Except ring extension 1/5

First, in a screening session, we instructed the participants to attempt performing various finger or wrist movements while recording high-density surface EMG (HDsEMG) signals from their forearms using three 64-electrode arrays (8×8; inter-electrode distance, 10 mm) (Fig. 1). Based on this screening, we selected for each participant one attempted movement that elicited discernible EMG but minimal observable movement that they would use for the one-dimensional control experiments. The selected attempted movements were thumb adduction, wrist abduction, and wrist abduction for P1, P2, and P3, respectively (e.g., Suppl. Video 1).

**Figure 1.**
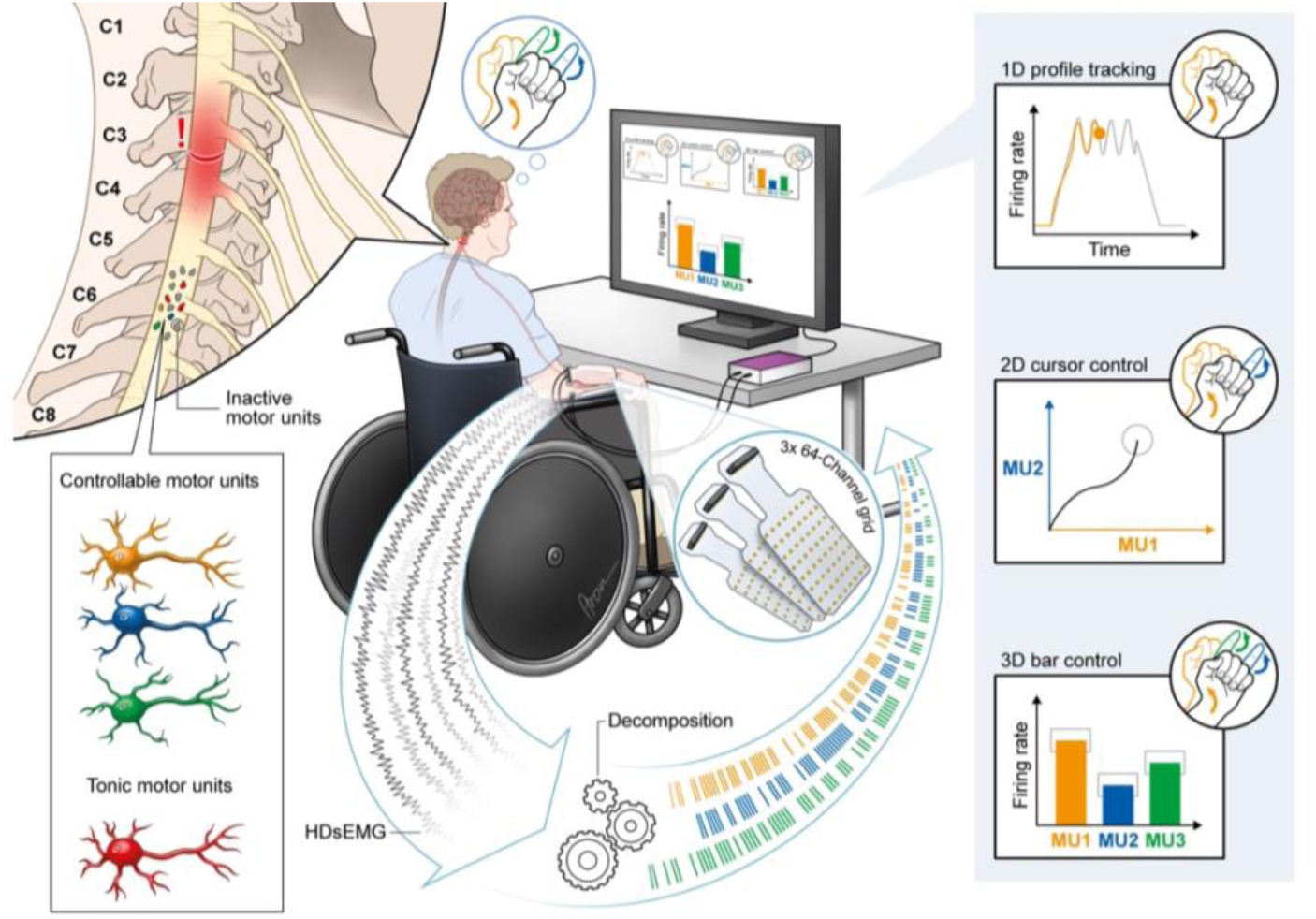
Investigating motor unit control in individuals with no hand function following a cervical SCI. Motor unit activity was detected through decomposition of high-density surface EMG (HDsEMG) recordings. We detected both “controllable motor units” that participants could modulate voluntarily, and “tonic motor units” that were not under volitional control, likely due to spasticity. Each colour (orange, blue and green) represents a different motor unit that could be independently controlled. Participants achieved flexible control for one-dimensional profile tracking, two-dimensional cursor control, and three-dimensional control of three different bars. This control was also extended to two different games, and simulated wheelchair navigation using three independent control signals.

To detect the activity of single motor units, at the beginning of each session we recorded HDsEMG signals as participants performed and sustained their selected attempted movement by controlling the global EMG amplitude to track a trapezoidal profile. We then used validated blind-source separation techniques^25,26^ to isolate the activity of single motor units from these HDsEMG recordings. For the online experiments, we applied the motor unit filters obtained through offline blind-source separation to the HDsEMG data in real-time. This allowed us first to visualise the spiking of the detected motor units as participants attempted their selected movements, and then to map them onto control signals for the online control experiments. Overall, we detected an average of 6.9 ± 3.1 (mean ± s.d., range, 2–12) motor units for the attempted movement across all participants and sessions.

Motor units could be classified into one of two distinct categories based on their online firing behaviour: controllable (62.5% of the units across all the recordings), and tonic (37.5 %) (examples in Fig. 2a, Suppl. Fig. 1). Controllable motor units were characterised by activity exclusively during movement attempts, while tonic motor units fired continuously during both rest and movement attempts, likely due to underlying spasticity^21,22^. Our prediction was that participants would be able to perform the tasks well using these controllable motor units, even in the presence of tonically firing units.

**Figure 2.**
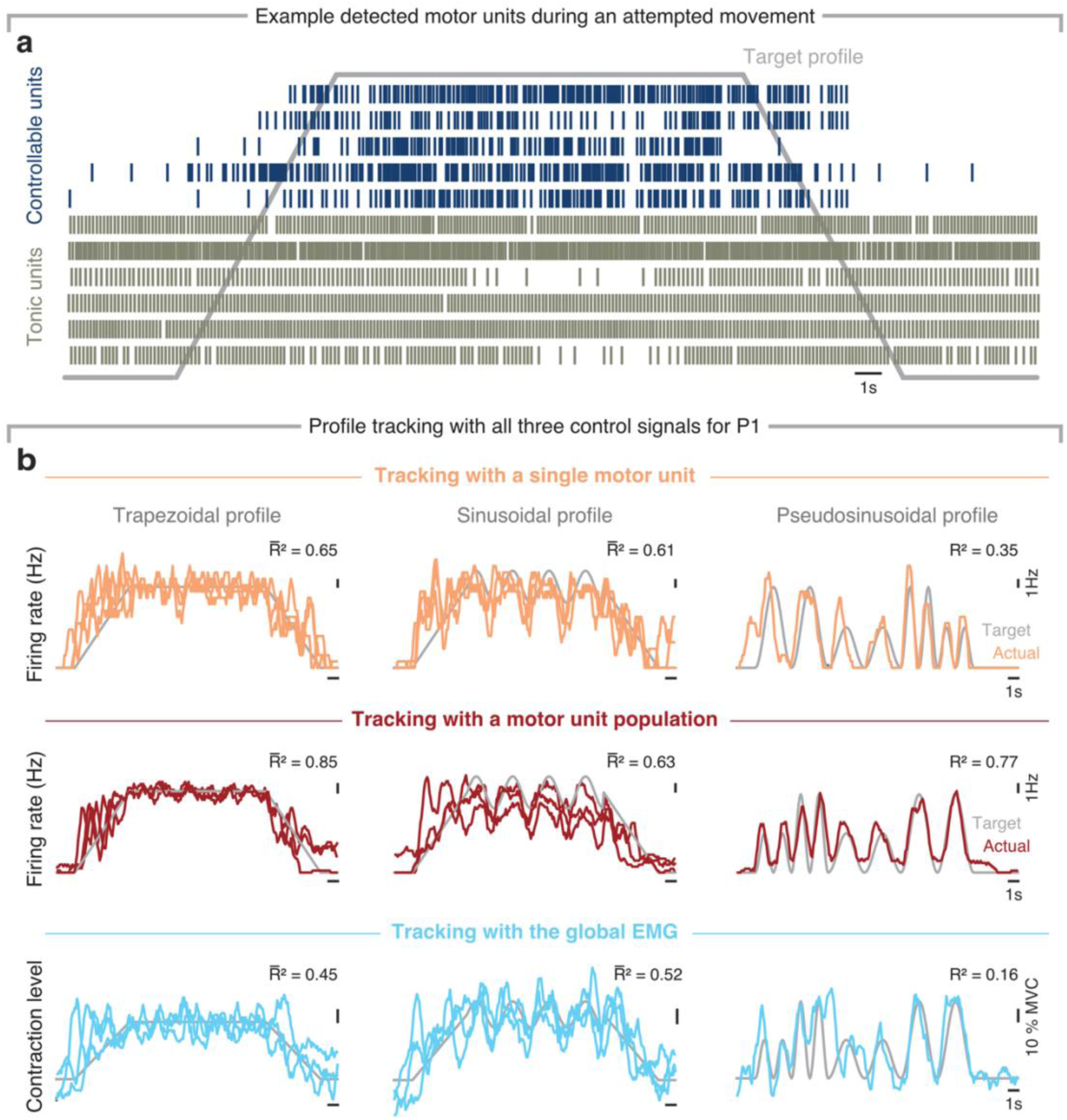
Intuitive motor unit control on the first day. **a.** Example detected motor unit activity, showing a representative combination of controllable units, whose activity could be volitionally modulated, and tonically active units likely related to spasticity. Data from P1. **b.** Online control for the first session from P1 as he tracked the three profiles using the activity of a single motor unit, a population of motor units, or the global EMG. Tracking accuracy was measured with the coefficient of determination (*R*^2^); *R^-^*^2^ is the average tracking accuracy across multiple trials. Each pseudosinusoidal trial was unique, so only one trial is presented for clarity. Tracking for P2 and P3 are shown in Suppl. Fig. 2.

### Motor unit activity enables intuitive and accurate proportional control across different one-dimensional tasks

We first investigated whether participants could harness the activity of identified motor units for precise continuous control by asking them to control the vertical position of a cursor. We designed three types of target profiles, each imposing a different control requirement (Fig. 2b): a trapezoidal profile, which required maintaining the control signal at a constant level, similar to holding a posture; a sinusoidal profile, which required smooth and predictable modulation; and a pseudosinusoidal profile, which demanded flexible control of both timing and intensity. For motor unit-based control, we tested both the participant’s ability to modulate the firing rate of a single motor unit, or of a population of motor units associated with the same attempted movement —often referred to as a cumulative spike train^27^ (CST)—, to better capture the overall behaviour of all active spinal motoneurons in a pool^28^. To test our prediction that motor unit activity should provide better control than the global EMG, participants also repeated all the profiles using the global EMG amplitude as control signal.

All participants achieved stable proportional control across the three target profiles from the first session (Fig. 2b, Suppl. Fig. 2, Suppl. Video 1). For P1 and P2, control based on a population of motor units yielded the highest accuracy, followed by single motor unit control; EMG-based control performed the worst (Fig. 2b and Suppl. Fig. 2a). For P3, EMG-based control was as accurate as for a population of motor units in the trapezoidal and sinusoidal profiles, and performed best in the pseudosinusoidal profile (Suppl. Fig. 2b); we attributed this to the surface EMG being relatively sparse and thus strongly resembling his detected motor unit population (Suppl. Fig. 3). Yet, overall, control of a population of motor units demonstrated excellent accuracy, outperforming all other modalities in virtually all comparisons. Crucially, such accurate control was not driven by overt movement, as demonstrated by the very low correlation (<0.3) between this pooled activity and any flickers of movement (Suppl. Fig. 4).

### Motor unit control is immediately intuitive, improving only modestly with practice

We next assessed whether participants could improve their control with practice. Participants tracked the three profiles using motor units associated with the same attempted movement for four to five sessions spanning a period of four to twelve weeks (examples in Fig. 3a). We noted a modest improvement in tracking accuracy that plateaued by the third or fourth session (Fig. 3b, Suppl. Fig. 5,6). This improvement was not related to an increase in the number of motor units over sessions, which did not change significantly over time (Suppl. Fig. 7a), and happened despite the participants only occasionally controlling the same motor units on different sessions (Suppl. Fig. 7b,c). Combined, these observations suggest that the performance gains stemmed from increased familiarity with the control. Importantly, these gains seemed to hold over time: we tested P1 for retention by having him repeat these experiments more than 60 days after his last training session and found that tracking performance remained comparable to the final —not the first— training session (Suppl. Fig. 8).

**Figure 3.**
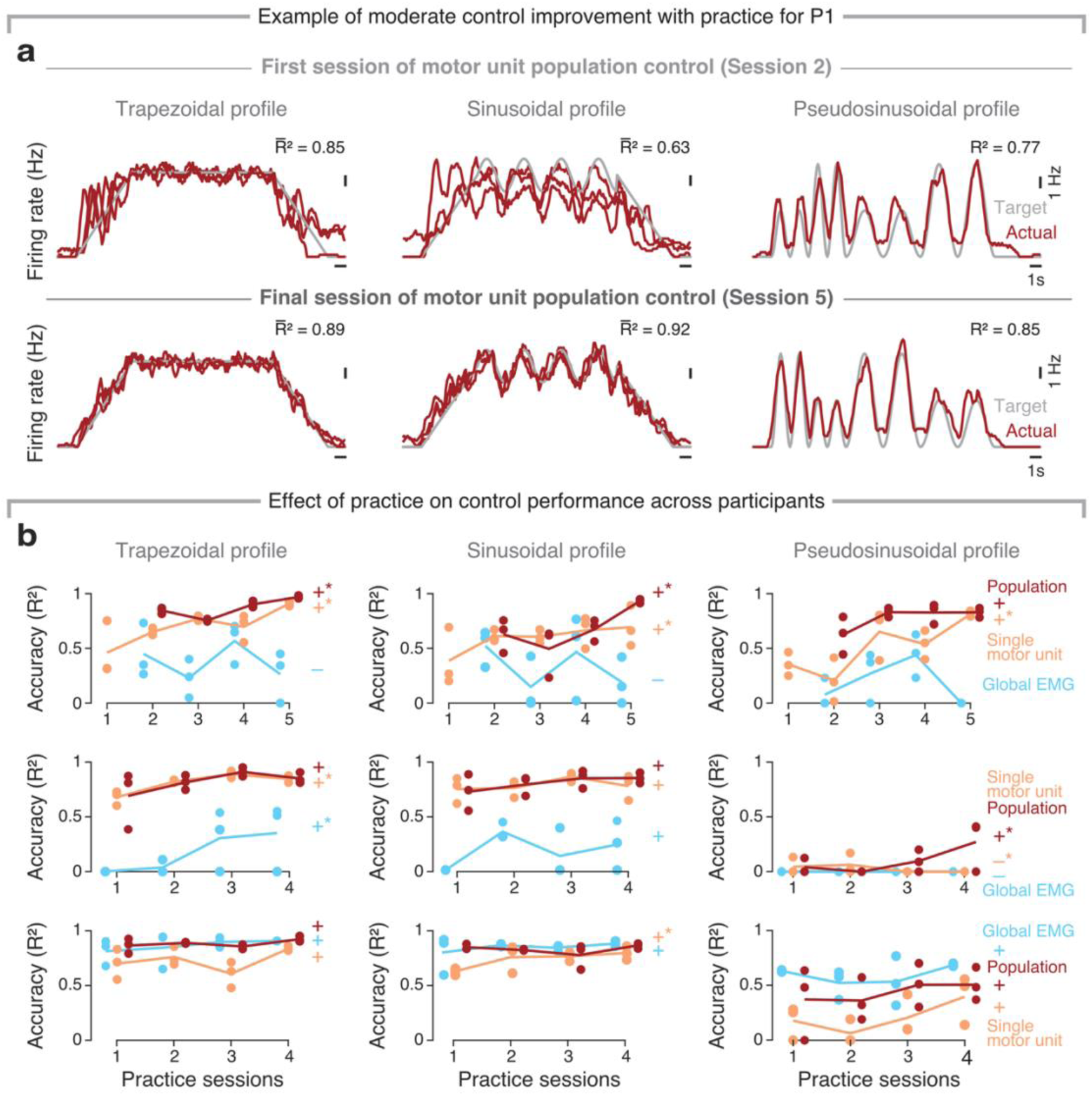
Motor unit control improves modestly over practice sessions. **a**. Example tracking for all three profiles and control signals for the first and last session from P1. **b**. Tracking performance across practice sessions spanning four to twelve weeks for all three participants. For P1, EMG and motor unit population control started from the second session. We fitted linear regressors to the across-day performance trends; + indicates a positive slope, - a negative slope, no sign a zero slope, * a statistically significant change (*P*<0.05, *t*-test); signs are colour-matched with the control signals.

The accurate tracking performance on the first session, along with the modest improvements over successive sessions, suggest that motor unit-based control was intuitive. To test this notion more directly, we assessed the control performance from motor units associated with attempted movements that did not undergo repeated practice (termed “untrained movements”: wrist flexion for P1, and little finger extension for P3—we were unable to test P2 due to time constraints). Consistent with the view that motor unit control is intuitive, participants could also accurately control motor units associated with these untrained attempted movements, with almost comparable performance as for the trained movements, even on their first session (Fig. 4).

**Figure 4.**
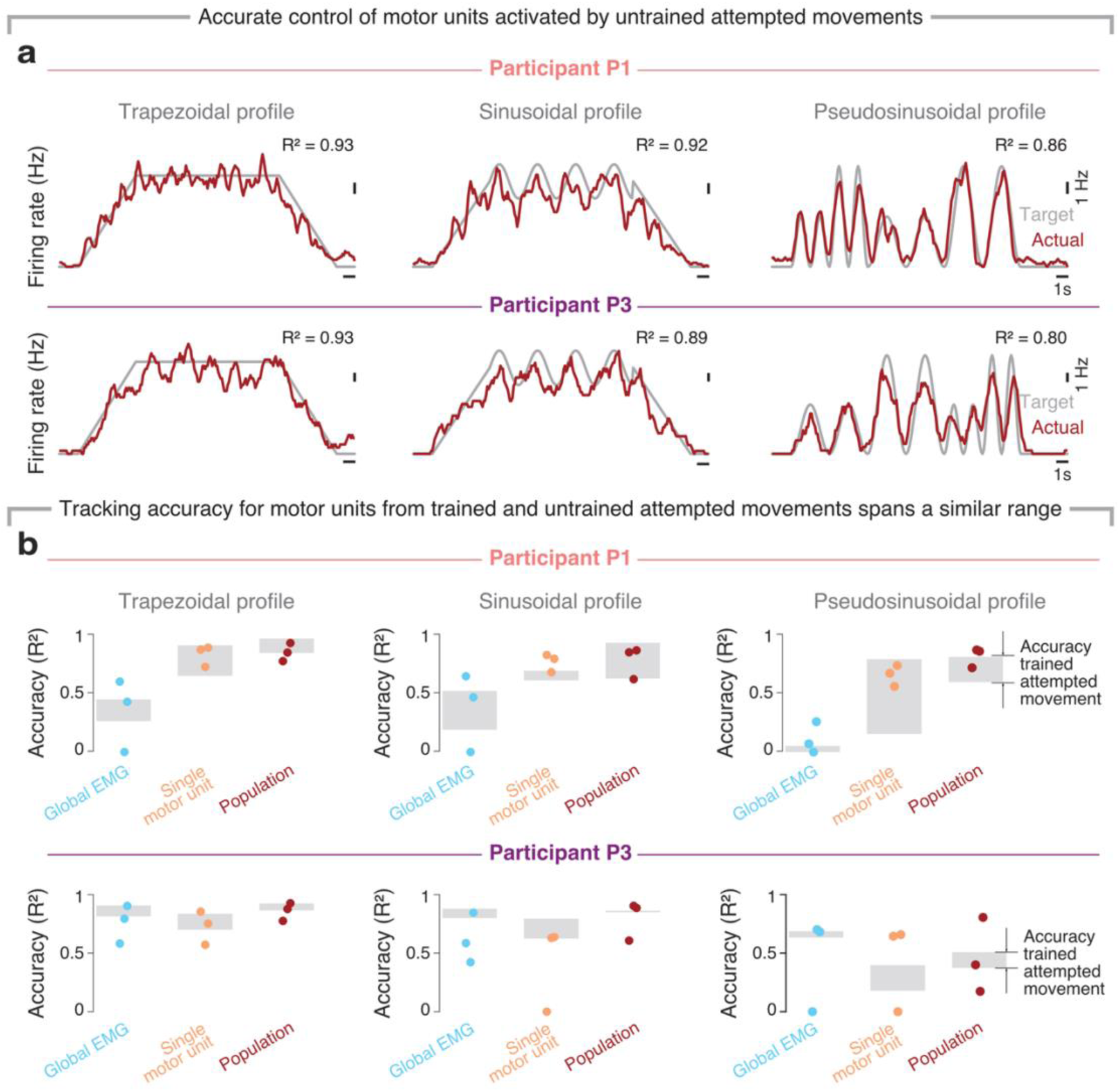
Accurate control with motor units associated with untrained attempted movements. **a**. Example tracking for all three profiles and control signals for untrained attempted movements for the last session from P1 and P3. **b**. Comparison of tracking performance for trained and untrained attempted movements. Untrained movements were tested for three sessions; participants completed one trial per session for each tracking condition (coloured circles). Grey rectangle, tracking performance for trained movements; bottom, tracking accuracy on the first session; top, tracking accuracy on the last session. Note that performance for trained and untrained spans a similar range for most comparisons.

### Flexible two-dimensional and three-dimensional control using multiple independent motor units

Restoring hand function demands more sophisticated control than the one-dimensional control described above. We therefore investigated the feasibility of controlling two independent motor units for cursor navigation in a two-dimensional workspace. We instructed participants to select two different attempted movements, to use one motor unit associated with each of them as a different control signal. P1 selected thumb adduction and wrist abduction, P2, wrist abduction and ring extension, and P3, thumb flexion and little finger extension. Then, in an initial calibration phase, we verified that the selected motor units could be activated independently (Methods).

Participants demonstrated competent two-dimensional cursor control immediately, being able to acquire 16 different targets per block and hold the cursor for >375 ms (Fig. 5a,b, Suppl. Video 2). Overall success rate was 85 %, 96.25 %, and 92.5 % for P1, P2 and P3, respectively, with an average time to success of 2.74 s, 4.08 s, and 4.67 s, respectively. Participants could easily acquire targets along either the horizontal or vertical axis, confirming their ability to independently modulate these motor units (Fig. 5a). For off-axis targets, participants found it easier to activate motor units sequentially rather than coactivating them, resulting in curved trajectories (Fig. 5a). Yet, this control was robust: for example, one of the participants exhibited myoclonic jerks that transiently hindered his control (Fig. 5a), but he could regain it immediately after and complete the trial.

**Figure 5.**
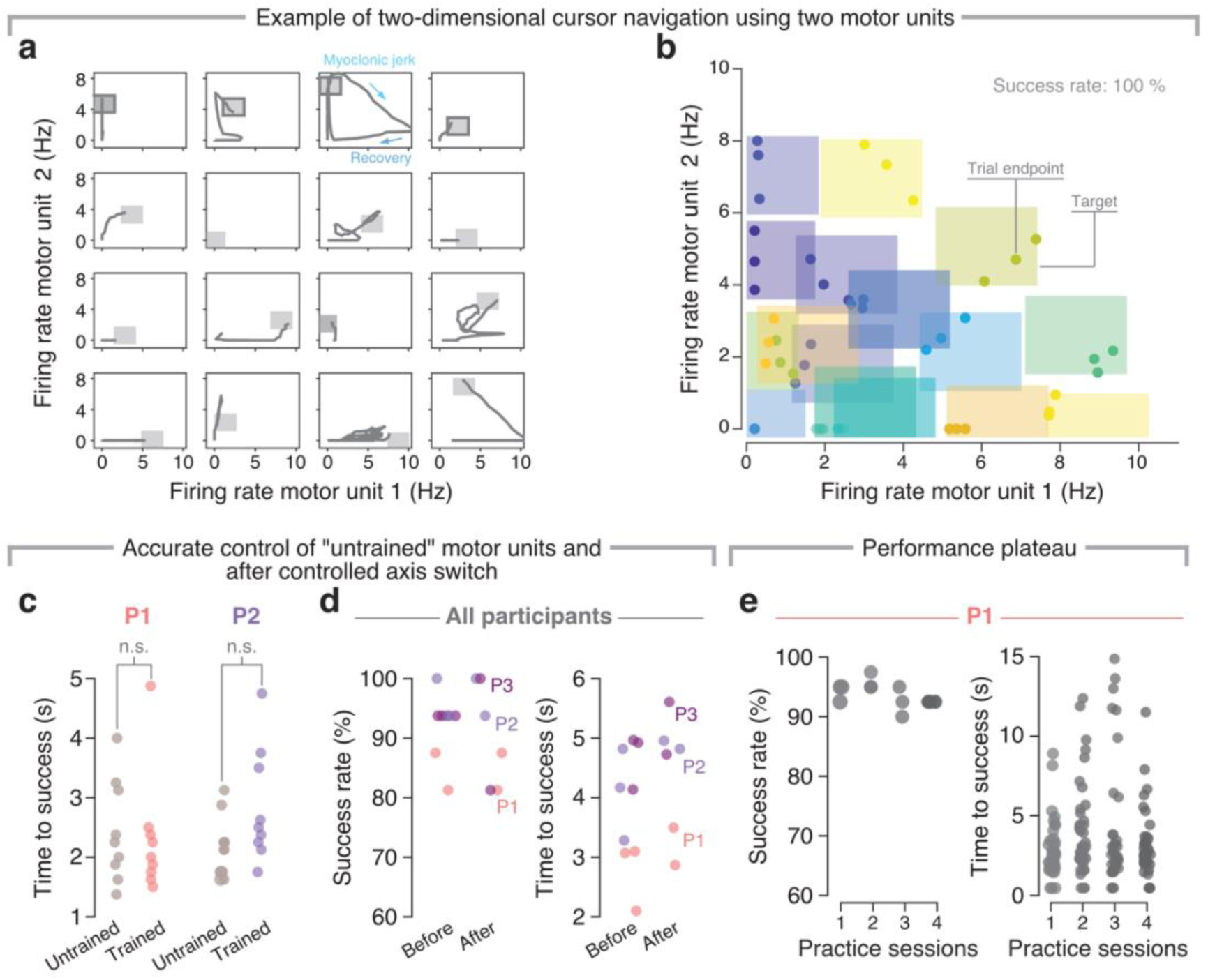
Two-dimensional cursor navigation using two independent motor units. **a**. Example cursor trajectories for sixteen trials from the same session from P3. During one trial the participant suffered a myoclonic jerk but recovered control quickly (shown with blue arrows). **b.** Example targets (coloured rectangles) and cursor position (coloured circles) at the end of the hold period for one session from P3. **c**. Performance during two-dimensional control was not significantly different for trained and untrained movements according to success rate and time to completion (*P*<0.05, paired *t*-test). **d**. Success rate and average time to trial completion for all participants were comparable before and after switching the mapping between motor unit firing rate and controlled axis. **e**, Control performance over four sessions of practice for P1 shows no strong trend.

Our two-dimensional control task design incorporated asymmetrical training histories: one motor unit was associated with an attempted movement that participants had extensively practiced during one-dimensional control (Fig. 3), while the other corresponded to a novel, untrained attempted movement. Based on our observation of similarly accurate one-dimensional control for motor units associated with attempted trained and untrained movements (Fig. 4), we also expected two-dimensional control performance to be similar across “trained” and “untrained” motor units. That is what we found: performance for targets on either the horizontal or vertical axis, which required the independent activation of the trained or untrained motor units, showed no noticeable differences (Fig. 5c). Moreover, P2 additionally performed two-dimensional cursor navigation with two “untrained” attempted movements (thumb extension and wrist extension) ∼9 months later, achieving comparable performance to when he used at least one “trained” movement (success rate, 95 %; average time to success, 6.94 s).

We further evaluated the intuitiveness of two-dimensional motor unit control by introducing an axis-switching manipulation in which the cursor control axes were swapped: motor unit one, initially controlling the horizontal axis, now controlled the vertical axis, and vice versa for motor unit two. Performance was not significantly affected by this new mapping: task success rate remained stable, and there was only a small increase in completion time (Fig. 5d). Finally, having observed similarly accurate control for trained and untrained motor units (Fig. 4), we tested the prediction that two-dimensional control would already be operating at near maximal performance from the first session and would not improve with practice. As expected, P1 showed no clear performance gains after five sessions (Fig. 5e).

Can participants control more degrees of freedom using motor units from paralysed hand and wrist muscles? To answer this, we explored three-dimensional control in P1 and P2 using a “three bars” task. After a calibration procedure like that for two-dimensional control, P1 selected motor units associated with attempted thumb adduction, wrist abduction, and wrist flexion, and P2 with wrist abduction, ring extension, and index extension. Both participants successfully achieved independent, proportional, real-time control over three motor units (Suppl. Video 3). This control again demonstrated notable flexibility: P1 adeptly performed the two-dimensional cursor control task using all possible pairwise combinations of the three independent motor units (Fig. 6a). Both participants were also able to selectively activate one motor unit to a target level whilst keeping the other two quiescent (Fig. 6b,c), but found simultaneous coactivation of multiple motor units more difficult (Fig. 6d,e), reminiscent of difficulties in two-dimensional control. Combined, these results show that although simultaneous control of multiple motor units is challenging, people with severe paralysis resulting from a cervical SCI can intuitively control multiple independent motor units from wrist and hand muscles with minimal practice.

**Figure 6.**
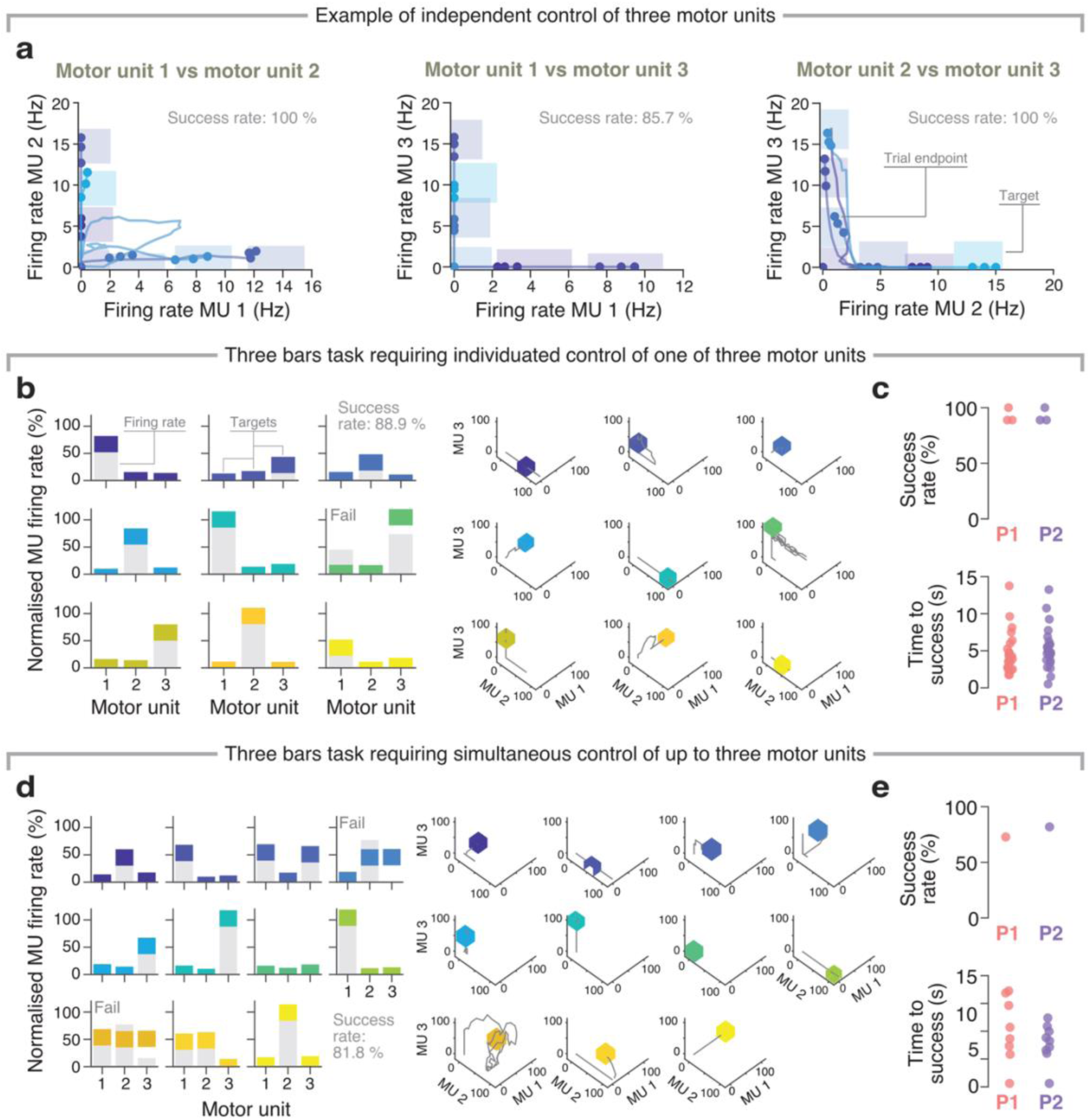
Independent and simultaneous control of three motor units. **a**. Pairwise tests of the independence of three motor units in the two-dimensional control task in one session from P1. **b**. Example trials from a three bars task that required isolated activation of one of three motor units; right panel, control trajectories in a three-dimensional normalised firing rate space. Coloured rectangles, target firing rates; grey bars, actual firing rates. Firing rates were normalised for visualisation. **c**. Performance metrics across all sessions for the task in panel b. **d**. Similar to b, but for a version of the three bars task that required simultaneous activation of multiple motor units. **e**, Performance metrics across all sessions for the task in d. MU, motor unit.

### Applications to gaming and assistive devices

We have so far focused on laboratory tasks designed to understand the ability of SCI participants to control the activity of motor units from paralysed muscles. Yet, successful deployment of this technology for assistive devices or even rehabilitation could benefit from extensive practice well beyond the relatively limited number of sessions that we have explored here. As a first step towards more engaging, gamified training, we have developed adapted versions of Pong^29^ and Snake^30^. P2 and P3 played Pong, controlling the vertical movement of the paddle by modulating the firing rate of either a single motor unit or a population of motor units (Fig. 7a, Suppl. Video 4). P1 and P3 played Snake, where the activation of one motor unit made the snake turn 90° clockwise, and the activation of another, 90° counterclockwise (Fig. 7b, Suppl. Video 5). All participants demonstrated proficient control after an initial familiarisation period, although, for Snake, both participants cited difficulties with directional orientation, as the snake’s turning mechanics varied based on its current trajectory.

**Figure 7.**
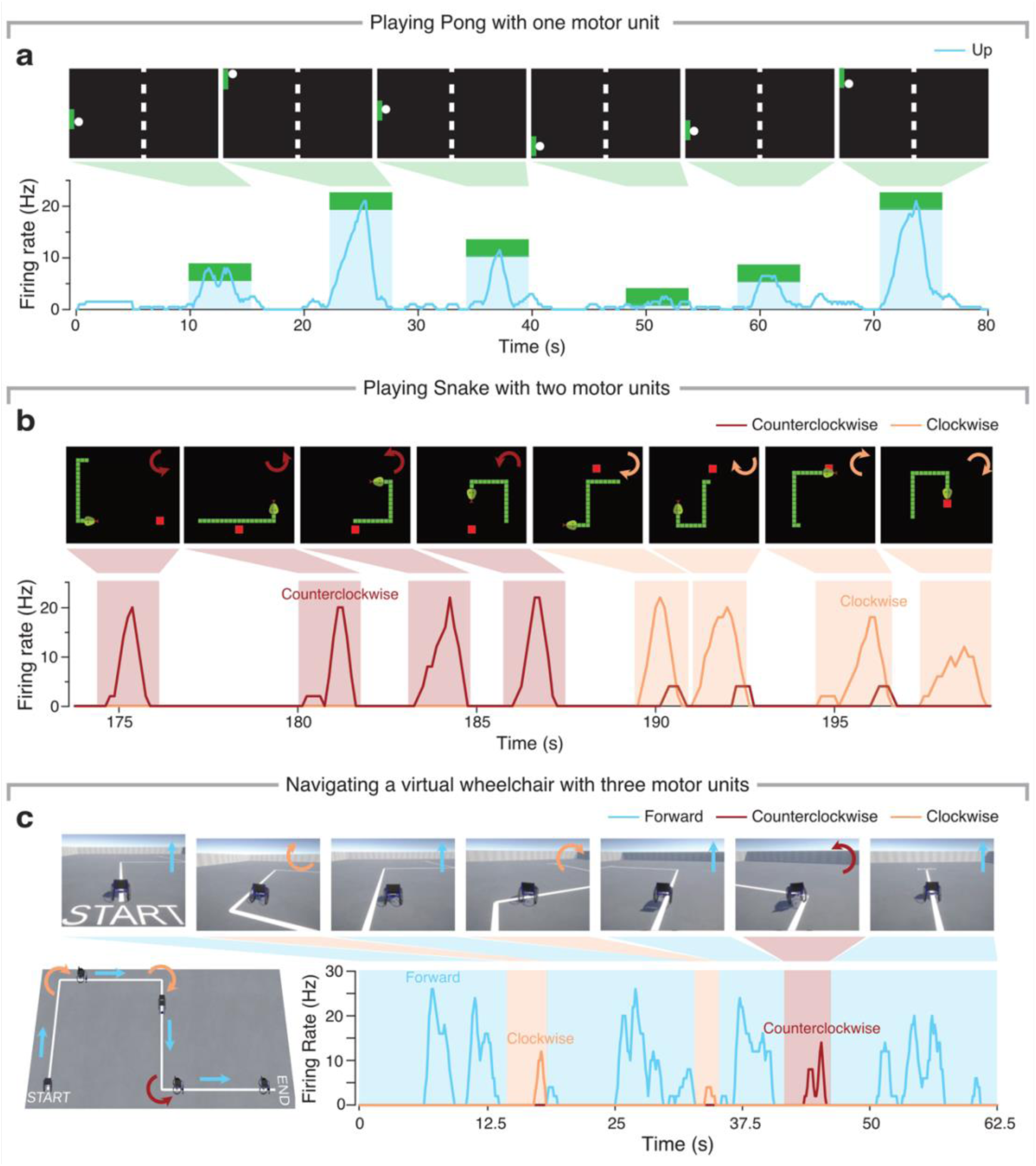
Playing games and controlling a (virtual) assistive device using the activity of up to three motor units. **a.** Playing Pong with one motor unit. Top, position of paddle and the ball during 80 s of play; bottom, firing rate of the motor unit controlling the paddle position; blue trace, motor unit firing rate; green rectangle, position of the paddle. **b.** Playing Snake with two motor units. Participants had to bring the snake (in green) to its food (in red). Top, example position of the snake and the food during ∼25 s of play; bottom, firing rate of the two motor units used for control. **c.** Independent control of three motor units to drive a virtual wheelchair along a target path towards a goal location. Top, example screenshots during a trial. Bottom left, target trajectory and example positions of the virtual wheelchair for one example trial. Bottom right, activation of the three motor units controlling the different actions.

In a step towards controlling an assistive device, we asked P1 to navigate a virtual wheelchair using the activity of three motor units (Fig. 7c), where one motor unit controlled the forward movement, and the other two the left and right turns. P1 could immediately navigate the wheelchair along the path with remarkable precision (Fig. 7c). Importantly, all participants expressed enthusiasm about using their residual motor unit activity for gameplay and (virtual) wheelchair control. This positive feedback underscores the potential of this technology to provide both engaging, recreational experiences and effective assistance for individuals with SCI.

## Discussion

We have demonstrated that individuals with no hand function following cervical SCI can harness the activity of motor units from paralysed muscles to achieve intuitive, continuous, and multi-dimensional control despite the lack of overt movement. Importantly, the controlled motor units were detected non-invasively through decomposition of high-density surface EMG recordings^25,31,32^, which we could accomplish even for a participant classified as motor complete, and for others who exhibited clinical signs of spasticity. These findings have direct implications for the development of assistive technologies for people with paralysis, and may pave the way for future rehabilitation interventions for spinal cord injury^33^ or stroke^34,35^.

### Both single and multidimensional motor unit control are intuitive

All three participants showed good one-dimensional control from the outset (Fig. 2b, Suppl. Fig. 2), with only small gains following several sessions of practice (Fig. 3). This suggests that, despite the severe injury to the descending pathways projecting to spinal motoneurons, motor unit control remains inherently intuitive, likely because it builds on brain structures that remain unaltered^36^. Consistent with this view, controlling motor units associated with attempted movements not explicitly trained was similarly accurate from the outset (Fig. 4), and a participant re-tested after two months showed no performance loss (Suppl. Fig. 8). This immediately intuitive control contrasts with “neurofeedback paradigms” requiring extensive training^37–39^, and supports the application of motor unit control for assistive devices.

Even if participants could readily expand their accurate control to two or three degrees of freedom, our work reveals looming challenges. During cursor navigation in a two-dimensional workspace, targets off the horizontal and vertical axes, which required coactivation, were more difficult to reach (Fig. 4). Indeed, participants tended to activate motor units sequentially, producing curved trajectories. This difficulty to coactivate may be because the combination of attempted movements was often unnatural (for example thumb flexion and little finger extension for P2), and hence it may have been difficult to control the attempted contraction of those muscles simultaneously^40^. In future work, we will select movements that are naturally co-occurring and hence more intuitive to coactivate.

We noted that some participants had difficulty holding the cursor at a particular location for long periods of time (Fig. 5b). An interesting way to simplify this problem would be to change the nature of the mapping between motor unit firing rate and cursor behaviour. For instance, instead of using a position mapping, we could use a velocity-mapping, as it is often done in intracortical BCIs^41,42^. Other alternatives would include Kalman filters^43^ machine learning techniques^44,45^, or shared human-machine control^38^, which would further reduce the cognitive load for the user. Even simple optimisation of the scaling of the control mapping could lead to important improvements in the usability beyond the proof of concept provided in this study.

### EMG decomposition into constituent motor units improves control

Isolating the activity of single motor units improved control relative to the global surface EMG by enhancing the fidelity of the control signal. This was due to two reasons. First, HDsEMG decomposition filters out non-physiological changes that are intrinsic to surface EMG recordings^46^, for example, due to fluctuations in skin conductance^47^. In contrast, these changes imposed the need for frequent EMG baseline recalibration during the experiment —changes that were not necessary for motor unit control, which remained unaffected. Second, we could exclude unwanted motor units as control signals, which proved crucial due to the presence of a large number (∼35 %, Suppl. Fig. 7a) of tonically active motor units, likely due to spasticity^21,22^. Such filtering is not possible for the global EMG^46^, which reflects the summed contributions of all motor units indiscriminately^48^. Crucially, offline decomposition alone was not enough to classify motor units as tonically firing or controllable; only real-time tracking confirmed which neurons were under voluntary control.

While control based on both populations of motor units and single motor units was very accurate, even without any additional postprocessing^49^, motor unit populations tended to outperform single motor unit control. This may be attributed to the inherent nonlinear response characteristics of motor units, which makes it difficult to control the firing rate of a specific motor unit to their maximum value over a long period of time^50^. Pooling multiple motor units together may average out these naturally occurring fluctuations, explaining the slightly improved control.

### Limitations and future directions

Fatigue was a limiting factor for some participants, especially when maximal motor unit discharge rates were required. Like improving control accuracy, this is another challenge that could be addressed by exploring the alternative mappings discussed earlier. Activating independent motor units simultaneously was notably difficult but is crucial for complex control. In addition to asking participants to use more naturally occurring combinations of actions, future work could also explore if more natural control signals could be formed from functionally coactive populations of motor units^51^, as a biologically grounded approach to improve multidimensional control.

This work opens further promising avenues for future work. One involves sampling from more forearm and hand muscles, including intrinsic hand muscles, which may provide expanded control options^34^. Future paradigms might also benefit from more intuitive mappings, such as aligning cursor direction with anatomical movement patterns (e.g., wrist flexion with the arm pronated moves the cursor downwards), to reduce cognitive load. Expanding the scope of application into real-world settings is an exciting opportunity, e.g., by interfacing with consumer electronics. Studies comparing motor unit control with standard assistive technologies (such as sip-and-puff systems or eye-tracking interfaces) will also be needed for translation into the real-world.

## Conclusion

This study provides evidence that individuals with lost hand function due to severe SCI can volitionally control the activity of motor units from paralysed muscles, which we detected with non-invasive recordings. This control was accurate, intuitive, and multidimensional, and could be achieved despite the presence of complete paralysis and spasticity. While we focused on individuals with cervical SCI, our approach is likely applicable to other motor-impaired populations, including those affected by stroke, peripheral neuropathies or myopathies. Some of these populations may benefit from using motor unit control not only for functional compensation, but also as a potential intervention to regain hand use.

## Methods

### Participants

This study was approved by the NHS HRA (REC reference 23/SW/0140) and Imperial College Research Ethics Committees (ICREC reference 19IC5641) and performed in accordance with the Declaration of Helsinki.

Three male participants (P1, P2, P3) who suffered from a spinal cord injury around C1 to C4 level were included in this study. Each participant provided written informed consent prior to participation. P1 is a male in his 30’s who suffered a level C4 injury 24 months before the experiment. He was clinically classified as AISA C (motor incomplete injury) and could generate no functional movements with his left hand and wrist (MRC power 0-1 in hand and wrist), except for the wrist extension. P2 is a male in his 60’s who suffered a spinal cord injury at C1-C3 level 14 months prior to participating in this experiment. He was classified as ASIA C (incomplete tetraplegia) and had no movement in his right upper limb, rated MRC grade 0/5, except for a flicker of movement in his ring finger, rated MRC grade 1/5. P3, a male in his 50’s who sustained a SCI at C2-C4 level 12 months prior to the experiment. He was classified as ASIA A (motor complete and had no functional movement in his both hands and power were rated MRC grade 0-1/5 in the hand and wrist. The patient characteristics are summarised in Table 1.

### Attempted movement screening and selection

In an initial calibration session, each participant was asked to attempt 16 movements (thumb flexion/extension/adduction/abduction, index flexion/extension, middle flexion/extension, ring flexion/extension, little flexion/extension, wrist flexion/extension/adduction/abduction) in turn while HDsEMG was recorded, and (flickers of) movement was observed. We identified candidate attempted movements as those where HDsEMG was observable while there was minimal observable movement. We asked participants to select from these candidate movements going forward.

### High-density EMG decomposition

We recorded HDsEMG signals using three 8 x 8 EMG grids (GR10MM0808, OT Bioelettronica, Turin, Italy) placed all around the forearm to cover all extrinsic wrist and hand muscles. The signals were recorded in a monopolar configuration (Quattrocento, OT Bioelettronica, Turin, Italy) using a customised software, sampled at 2048 Hz, and band-pass filtered (10–500 Hz). Each experimental session included a calibration part and an online control part. During calibration, the participant was instructed to track a trapezoidal profile with a maximum of 100 % (for P1) or 50 % attempted MVC (for P2 and P3) using visual feedback on the global EMG amplitude. Then, a validated blind source separation algorithm was applied to decompose the recorded HDsEMG signals into constituent motor unit activity using this calibration data^25,26^. The detected motor units were retained when the Silhouette (SIL) value was higher than 0.88; SIL is a clustering quality measure that reflects how well spike and noise centroids are separated after performing K-means clustering on extracted motor unit pulse trains. Manual editing was performed later to retain only high-quality motor units using standard approaches^52^. Duplicate motor units were detected by comparing their spike trains through cross-correlation. Each pair of motor unit spike trains was aligned across a ±50 ms lag range, and their similarity was measured as the fraction of spikes occurring within a ±5 ms coincidence window. Pairs showing more than 30% agreement were considered duplicates. For each duplicate set, the unit with the smallest inter-spike-interval coefficient of variation was kept. The identified motor unit filters for all the retained units were stored for later use for online visualisation and control.

Online estimation of motor unit spiking events was achieved by applying the stored motor unit filters to the incoming HDsEMG data^32,53^.Spikes were identified by measuring the Euclidean distance between each pulse train value and the spike and noise centroids derived from offline decomposition for each motor unit. Values nearer to the spike centroid were labelled as spikes. This method removed the requirement for real-time K-means clustering, and helped lower feedback latency. EMG signals were analysed in windows of 125 ms. Firing rates were calculated by counting the spikes detected over a 1000 ms sliding window.

We validated the accuracy of our online decomposition by comparing the motor unit action potential shapes (Suppl. Fig. 9a) and motor unit discharge times (Suppl. Fig. 9b) obtained through offline and online decomposition. In all cases we observed a very good match.

### Control signals

We compared the participants’ ability to control the activity of single motor units, populations of motor units associated with the same attempted movement, and the global surface EMG.

During calibration, the activity of individual motor unit was displayed in real time using a raster plot. Participants attempted their selected movements, and a single motor unit was selected for individual motor unit control based on its controllability determined by the investigators and the participant. Motor units that were not activated during the attempted movement and motor units that tonically fired non-discriminately during periods of contraction and rest were excluded. The firing rate of the remaining motor units (i.e., those under voluntary control) was averaged to form the motor unit population control signal. For control of the global surface EMG, bipolar EMG was first obtained by signal differencing along the muscle fibre direction; then, the overall EMG amplitude was calculated as the median RMS value of the EMG amplitudes across all channels from all grids. The RMS is the optimal estimator of EMG amplitude supposing the signal follows a Gaussian distribution^54^. All control signals were calculated in real-time over a 1000 ms window, sliding every 125 ms.

### One-dimensional tracking tasks

We designed three distinct tracking profiles to evaluate the participants’ ability to control their motor units subject to different task constraints, as well as to compare control signals (Fig. 2a). These were:

1. **Trapezoidal profile**: This profile increased linearly from 0 to the target MVC level within 6 s, remains constant for 15 s, and then decreased linearly back to 0 within 6 s.
2. **Sinusoidal profile**: This profile linearly ramped from 0 to the target MVC within 6 s, oscillated around the target MVC at an amplitude of ±20% of target MVC and a frequency of 0.25 Hz for 15 s, and finally ramped linearly back to 0 within 6 s.
3. **Pseudosinusoidal profile**: This profile consists of four sinusoidal segments, each spanning two cycles and possessing a frequency of either 0.25 Hz or 0.5 Hz and an amplitude of either 50% or 100% of the target MVC. The segments were randomly ordered (without repetition), phase-shifted backward by one-quarter of a cycle, and amplitude-shifted upward to ensure non-negative values. This procedure produced a seamless final profile.

The target MVC was 100% MVC for P1 and 50% MVC for P2 and P3. The tracking performance for each profile was assessed with the coefficient of determination (*R*^2^). Participants completed four or five sessions of one-dimensional control, on each day completing at least three trials of each of the three profiles using each of the three control signals indicated above (i.e., at least 27 trials per day).

The hand movements were recorded with a camera during the experiment. To quantify the hand kinematics during the task, we calculated the first principal component of pixel changes in the video during the tracking task, assuming that the changes of video pixels were mainly due to the 1D hand movements (Suppl. Fig. 3). The correlation between the principal component of hand kinematics and motor unit activity was then quantified with the *R*^2^.

### Two-dimensional cursor navigation task

We next assessed whether participants could control a cursor in two-dimensions using the activity of two independent motor units. We first stacked motor unit filters from two different movements to expand the dictionary of available motor units. Since each movement is performed separately, the offline HDsEMG per movement is associated with movement-specific whitening matrices. To account for statistical variability of the two different movements, new whitening matrix derived from the concatenated EMG of the two different movement selected was calculated. This permitted recalibration of the spike and noise centroids necessary for stable online decomposition across both movements. Duplicate motor units across grids associated with the different movements were additionally removed, using the same spike-based threshold detailed under “High-density EMG decomposition”.

Prior to the control experiments, we visualized the activity of all motor units using a real-time raster plot, and the screened for units with independence. A motor unit was considered independent if it was selectively activated by one attempted movement without concurrent activation from the others. Once two motor units were confirmed as being independent, one was assigned to the cursor’s position along the horizontal axis, and the other to the position along the vertical axis.

A two-dimensional cursor navigation task was designed to assess the control performance (Fig. 5a). Targets were distributed on the two-dimensional plane in a polar coordinate mode, with radial distances of 1/3, 2/3, and 1 of the maximum control range, and angular positions of 0, π/8, π/4, 3π/8, π/2, yielding a total of 16 targets. This “rainbow” target distribution was designed to avoid two motor units being activated to their maximum discharge rate the same time. Each target’s position was randomly jittered within ±5% of the maximum control range of both axes to introduce additional spatial variability. A tolerance range of ±15% of the maximum control range was set for both axes to define successful target acquisition (i.e., to define target size). In each session, participants were presented all of the targets in random order. A trial was considered successful if the participant acquired the target and maintained the cursor within its tolerance region for at least 375 ms. The 375ms holding period, corresponding to three consecutive EMG processing windows, was introduced to promote intentional control, rather than transiently passing through the target bounds. Maximum trial duration was 15 s; inter-trial time —either following a successful trial or a timed-out trial— was 5 s, to allow for a rest period. The maximum control range was defined as the average firing rate of the motor unit calculated from the offline decomposition of the trapezoidal profile tracking.

In each two-dimensional control experiment, participants were asked to complete three blocks of trials of this task. Subsequently, the control axes were switched to probe for generalisation of control across control axes: motor unit 1, which was initially used to control the horizontal axis, was used to control the vertical axis, and vice versa for motor unit 2. Participants were asked to complete another two blocks of trials after axis switch. Performance on this task was quantified based on task success rate and time to success.

### Three-bars task

Next, we expanded our approach for two-dimensional motor unit control to explore the feasibility of controlling three independent motor units. Like for two-dimensional control, we sought to identify an additional independent motor unit by decomposing HDsEMG signals from a third attempted movement. To validate the participant’s ability to independently control the three motor units, we first performed the two-dimensional cursor navigation task using pairwise combinations of the three motor units, restricting the targets to orthogonal axes and excluding diagonal targets.

In addition to this pairwise assessment, we designed a “three bars” task to more directly evaluate three-dimensional control. In this task, three targets were assigned to each motor unit, corresponding to 1/3, 2/3, and their full maximum firing rate, respectively. Here, maximum firing rate was again calculated as the average firing rate calculated from the offline decomposition of the trapezoidal profile tracking. A tolerance of ±15% of the maximum firing rate was allowed for successful target acquisition (i.e., to define target size). We designed two versions of this task: one to probe independent control, the other, simultaneous control. The independent control version of the task had nine conditions; in each condition, one motor unit was assigned to one of the three contraction levels while the other two remained quiescent (Fig. 5b). The participants were asked to complete three blocks of this task. The simultaneous control version of the task required not only individuation of single motor units, but also the coactivation of motor unit pairs. Three targets were assigned to each motor unit, corresponding to 0, 1/2, and the full maximum firing rate. Only coactivation with 1/2 maximum firing rate was considered to avoid simultaneous activation of multiple motor units to the maximum level. This led to 11 task conditions (Fig. 5d); participants were asked to complete one block of this coactivation bar task. The control performance of the three bars task was also quantified based on success rate and time to success.

### Pong, Snake, and virtual wheelchair control

To expand control outside of constrained laboratory tasks designed to probe performance and generalisation, we built virtual environments for participants to play adapted versions of Pong, Snake, and control a virtual wheelchair.

In Pong, participants controlled the vertical position of a paddle (1/5 of the game window’s height) to intercept a ball bouncing within the screen’s boundaries. The paddle’s vertical position was mapped linearly in real time to the firing rate of a selected motor unit or a population of motor units, such that higher firing rates produced greater upward displacement. This position was updated every 125 ms. Ball trajectories were semi-randomised and followed the law of reflection: each reset positioned the ball at the centre with a trajectory directed away from the paddle. The selected attempted movements for each participant were the same as those in the one-dimensional tracking.

In Snake, participants used two motor units to control 90-degree clockwise and counterclockwise rotations of a virtual snake within an 80×80 grid. The selected attempted movements for each participant were the same as those in the two-dimensional cursor navigation. The snake moved one step forwards every 330 ms, and participants could update the control commands every 125 ms. Participants had to select the correct motor unit at the right time to direct the snake towards food (red box), which appeared at random locations in the workspace. Once the snake entered the food’s area (with a tolerance of ±1 grid unit), the food was eaten, resulting in the snake’s overall length increasing by one. A trial was deemed to have failed if the snake had intercepted itself (rather than food); if the snake reached the end of the screen, it reappeared at the opposite side.

We finally rerouted the activity of three independent motor units that allowed P1 to drive a virtual wheelchair developed in Unity (Unity Technologies, San Francisco, USA). He controlled the forward, left turn and right turn of the wheelchair using thumb adduction, wrist abduction, and wrist flexion. The wheelchair moved one step for every 125 ms. P1 controlled the forward movement continuously by keeping activating the thumb adduction motor unit. The velocity of the wheelchair was proportional to the activation level of the motor unit. Discrete 90-degree turnings were achieved by activating the wrist abduction or wrist flexion motor units.

### Additional analyses

1. **Relationship between the global EMG and decomposed motor unit population activity** (Suppl. Fig. 3): To understand the relationship between the global EMG and the activity of the detected motor unit population, we calculated the Pearson’s correlation coefficient between the EMG RMS and the firing rate of the motor unit population during the trials in which participants tracked the different profiles using the global EMG. The motor unit activity was decomposed from the offline recorded HDsEMG using a pseudo-online mode, by applying the calibrated motor unit filters and parameters used during the online experiments. In addition, the EMG signal-to-noise ratio (SNR) was calculated to help elucidate the relationship between EMG “quality” and the activity from the detected motor unit population. The SNR was defined as the ratio of the RMS value of the global EMG signal during maximum voluntary contraction to the RMS value of baseline global EMG noise as 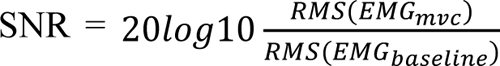
.
2. **Relationship between attempted residual movements and motor unit activity (**Suppl. Fig. 4): To quantify the relationship between motor unit population activity and the residual hand kinematics, we extracted hand kinematics from the recorded videos during the one-dimensional tracking tasks (Fig. 2). First, videos were cropped to hand-view frames and converted to grayscale. Subsequently, we performed a Principal Component Analysis of the image pixel value changes was calculated; we considered the first principal component of as an indicator of attempted movement. Finally, we measured the similarity between these inferred kinematics and the motor unit population activity as their coefficient of determination (*R*^2^).
3. **Compensation of control delays for pseudosinusoidal tracking** (Suppl. Fig. 5): For the pseudosinusoidal profile tracking, we noticed that participants could usually track the amplitude well but with apparent time delay. To better quantify the amplitude and time control, we segmented the full profile into eight semi-sinusoidal segments and then calculated the tracking accuracy after aligning the online profile to each segment using cross-correlation. The tracking accuracy after alignment and the estimated time delay with cross-correlation were then used to better characterize the pseudosinusoidal control.
4. **Offline validation of online motor unit decomposition** (Suppl. Fig. 9): To validate the accuracy of the online motor unit decomposition algorithm —beyond the ability of the participants to control these detected motor units—, we compared the motor unit action potentials and motor unit spike trains obtained from the online experiment with those from offline re-decomposition (i.e., offline decomposition of online recordings). Pearson’s correlation coefficient and rate of agreement were used to assess the correlation between motor unit action potentials and the consistency of motor unit spike trains between motor unit activity tracked online and verified offline, respectively.

### Statistics

The data were collected and analysed using MATLAB R2024b (MathWorks Inc.). The number of participants in this case series was not large enough for a robust inter-user statistical analysis, considering the large clinical diversity; therefore, results are reported for each participant individually.

To quantify whether continued practice improved one-dimensional motor unit control, we fitted within-participant linear regression models using the `fitlm` function. The sign of the fitting slope (β) was used to evaluate the effect of practice on control performance, and the *P* value, the statistical significance of the linear fit. *P* values were computed from the *t*-statistic (*t*), which assessed whether the slope (β) of the regression line was significantly different from zero^55^. To compare the performance for trained and untrained movements during the two-dimensional navitaion tasks, we first used a modified version of the Kolmogorov-Smirnov test after testing for normality using the `lillietest` function. If the data met the normality assumption, we used a parametric paired *t*-test (`ttest2` function) to assess statistical significance; otherwise, we used a nonparametric Mann-Whitney U test (`ranksum` function). This comparison was done within-participant by comparing performance across different trials. The statistical significance level was set to 0.05 throughout the paper.

## Data Availability

Data will be made available upon reasonable request to the corresponding author.

## Code availability

All the analysis will be made public upon publication in a peer reviewed journal.

## Acknowledgements

We thank our participants and their carers for their time, patience and efforts. We also thank the Alderbourne and Daniels Rehabilitation Unit at Hillingdon Hospital, including the clinicians, nurses and therapy staff for their assistance in identifying participants and providing us with the environment to conduct this study. Finally, we also thank Dr. Simon Avrillon for providing technical support and open-source software during the early stages of the project, and Dr. Balint Hodossy for contributing the virtual wheelchair environment. X.Y. is supported by the Fundamental Research Funds for the Central Universities, grant no. 2242025F20002. V.R. is supported by an NIHR Academic Clinical Fellowship. C.F.G. is supported by the Imperial College London President’s PhD Scholarship. This work was supported by the Imperial-META Wearable Neural Interfaces Research Centre (D.F. and J.A.G.), grant no. EP/T020970/1 from the UKRI Engineering and Physical Sciences Research Council (D.F. and J.A.G.), and grant no. ERC-2020-StG-949660 from the European Research Council (J.A.G.).

---

**Supplementary Figure 1.**
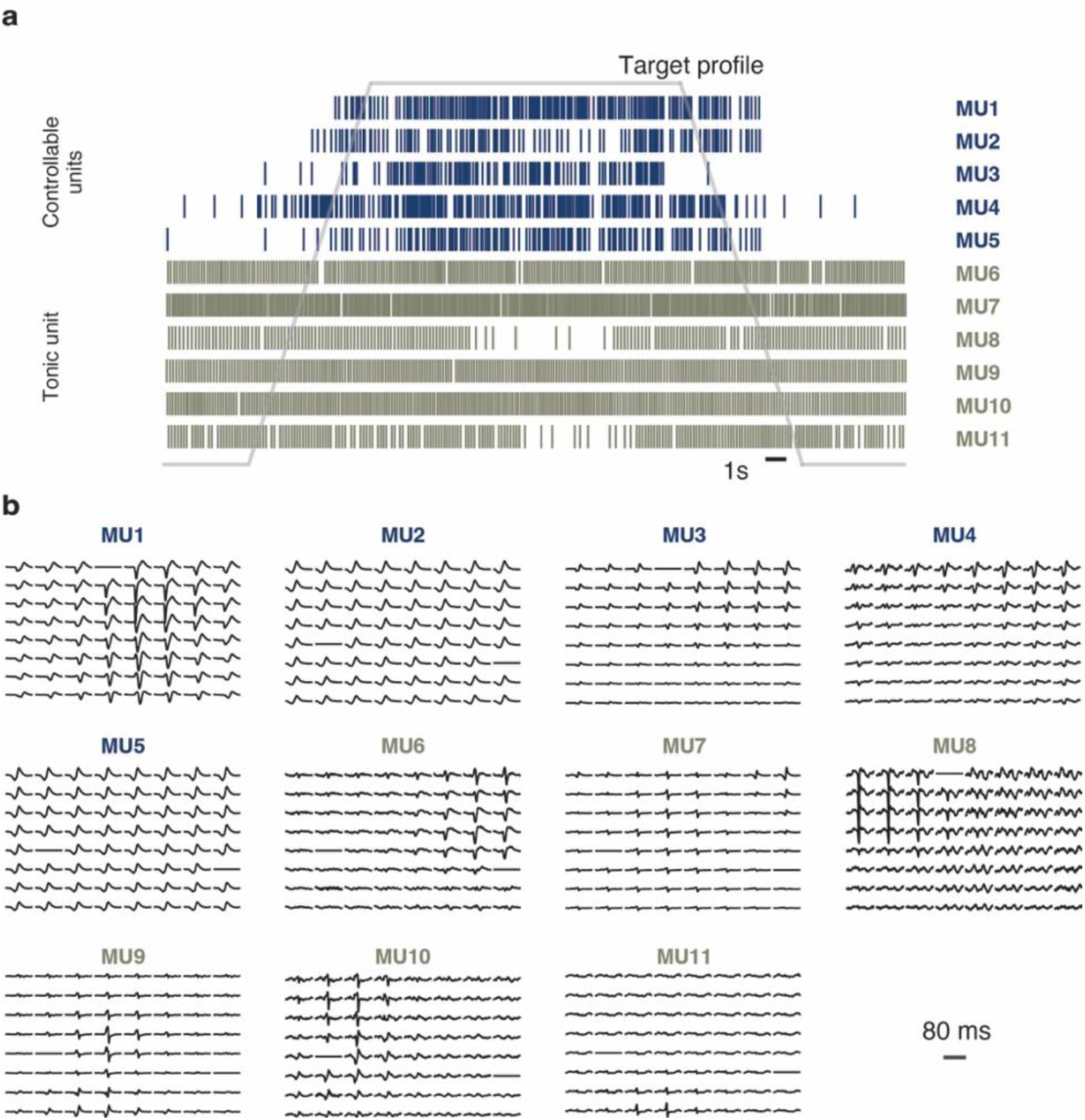
Motor unit action potential waveforms identified during one representative contraction. **a.** Example detected motor unit activity, showing a representative combination of controllable units whose activity could be volitionally modulated, and tonically active units likely related to spasticity. Data from P1, reproducing Figure 2a. **b.** Motor unit action potentials for the detected motor units across all 8×8 of one HDsEMG array. MU, motor unit.

**Supplementary Figure 2.**
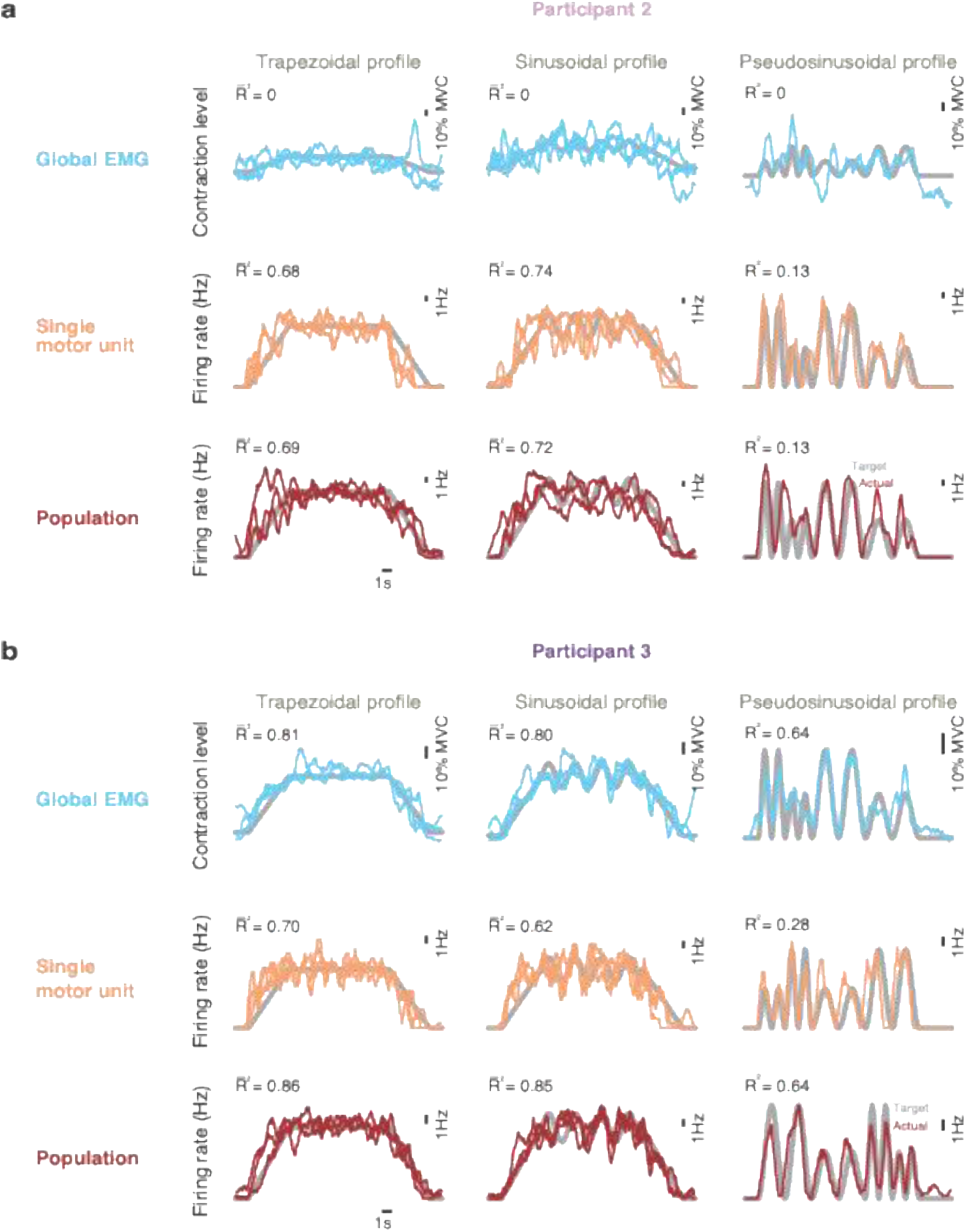
Intuitive motor unit control on the first day for P2 and P3. **a.** Control performance for the first session from P2 as he tracked the three profiles using the activity of single motor units, of a population of motor units, or the global EMG. Tracking accuracy was measured with the coefficient of determination (*R*^2^); *R^-^*^2^ is the average tracking accuracy across multiple trials. Each pseudosinusoidal trial was unique, so only one trial is presented for clarity. **b.** Same as above but for P3.

**Supplementary Figure 3.**
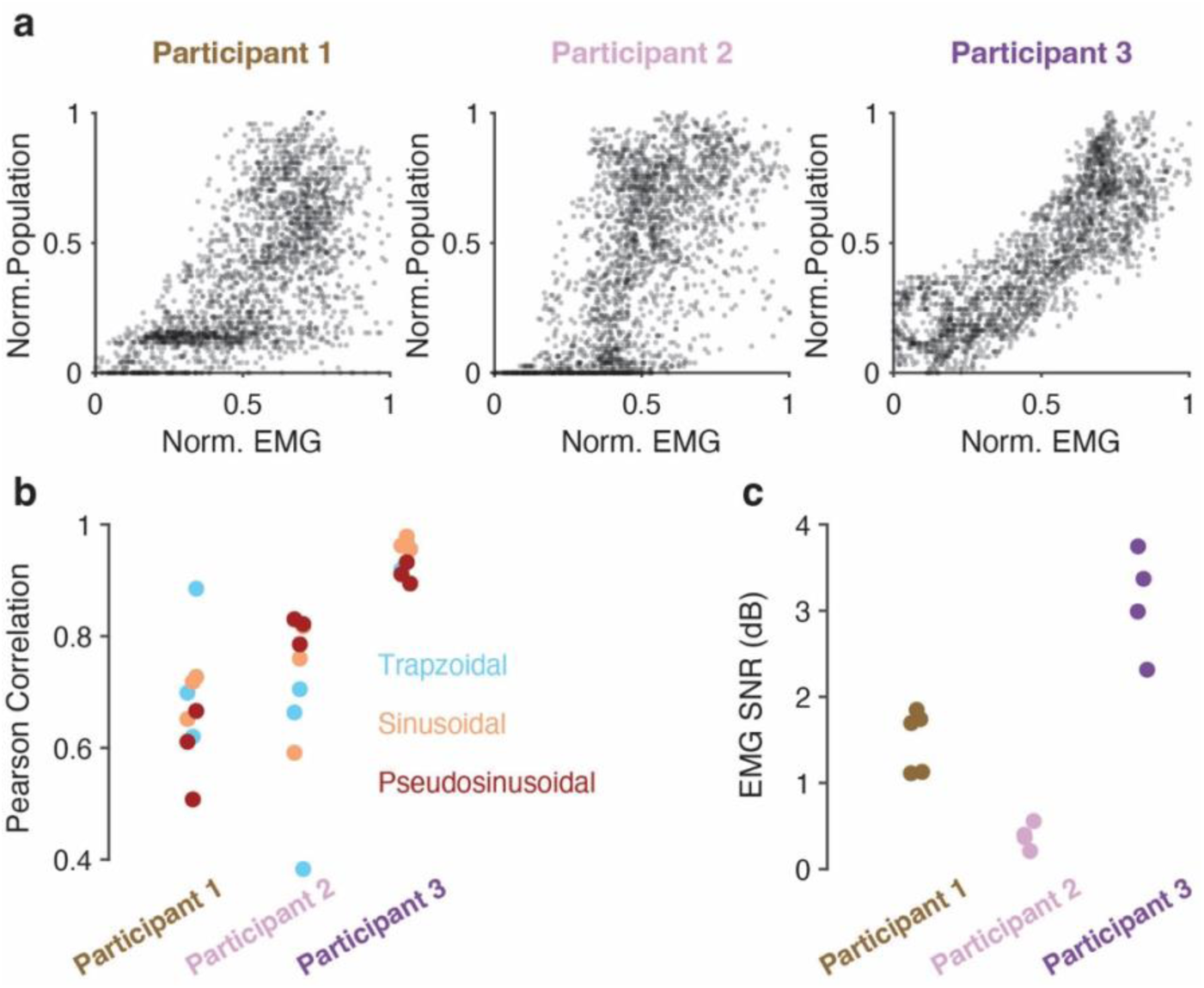
Assessing the relationship between the global EMG and motor unit population activity. **a.** Scatter plots showing the relationship between the global EMG and the motor unit population firing rates. Markers, all the time bins from all the trials from the first session using the global EMG for tracking. Note the stronger correlation for P3 compared to P1 and P2. **b.** Correlation between the global EMG and motor unit population firing rates for the three tracking tasks, measured based on panel a. Markers, individual trials from the first session. **c.** Signal-to-noise ratio (SNR) of the global EMG for each experiment session. Data points, individual sessions.

**Supplementary Figure 4.**
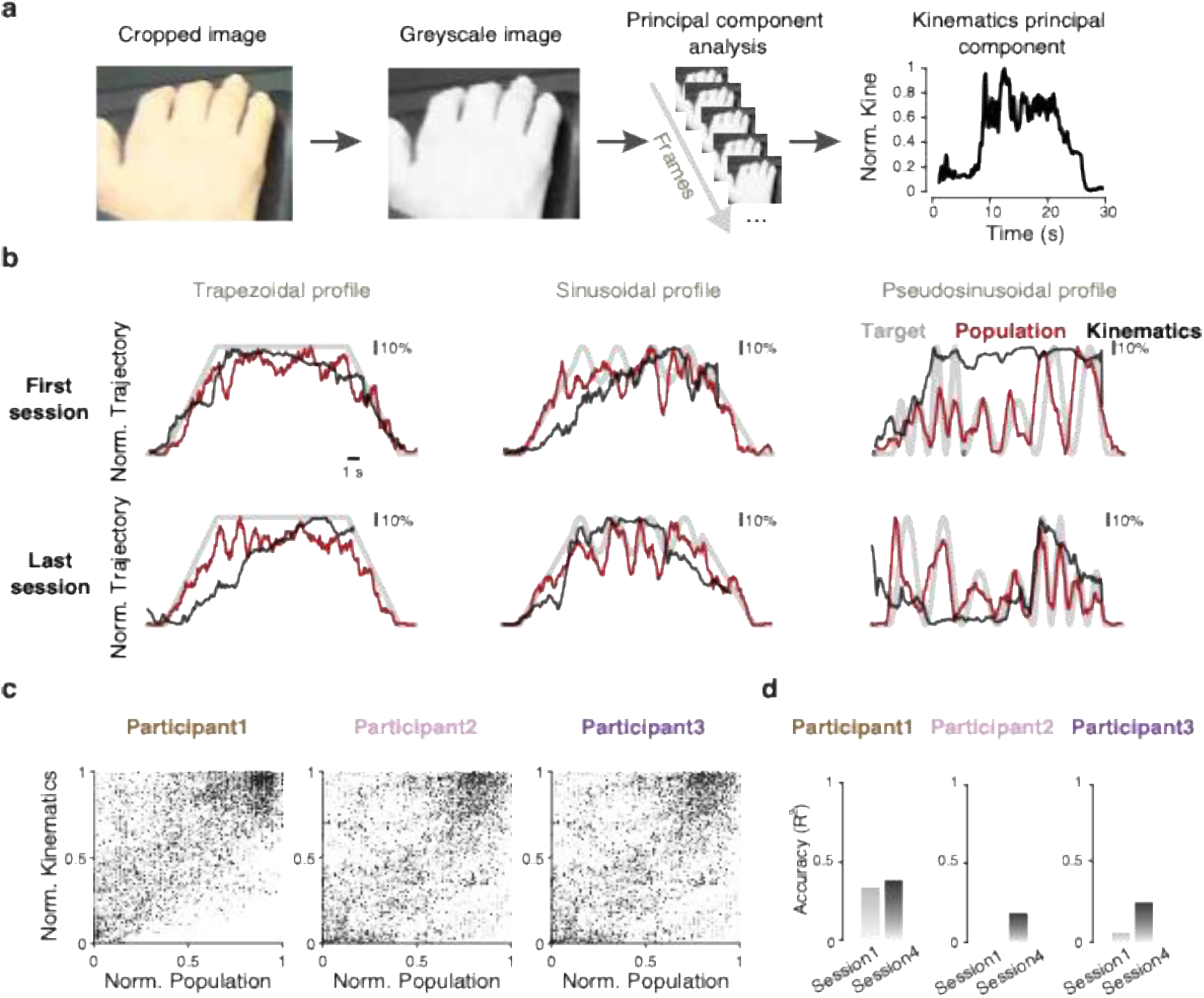
Lack of correlation between residual hand movements and motor unit activity during one-dimensional control. **a.** Pipeline to extract hand kinematics from the recorded videos as detailed in the Methods. **b**. Example inferred movements (black) and motor unit population firing rate (red) for the three profiles (grey) for P3. **c**. Scatter plots showing the lack of strong correlations between inferred hand movement and changes in motor unit population firing rate. Grey dots, data from session 1; black dot, data from the final session for each participant. **d.** Goodness of (*R*^2^) between the inferred hand movements and the motor unit population firing rate for the first and last sessions for each participant.

**Supplementary Figure 5.**
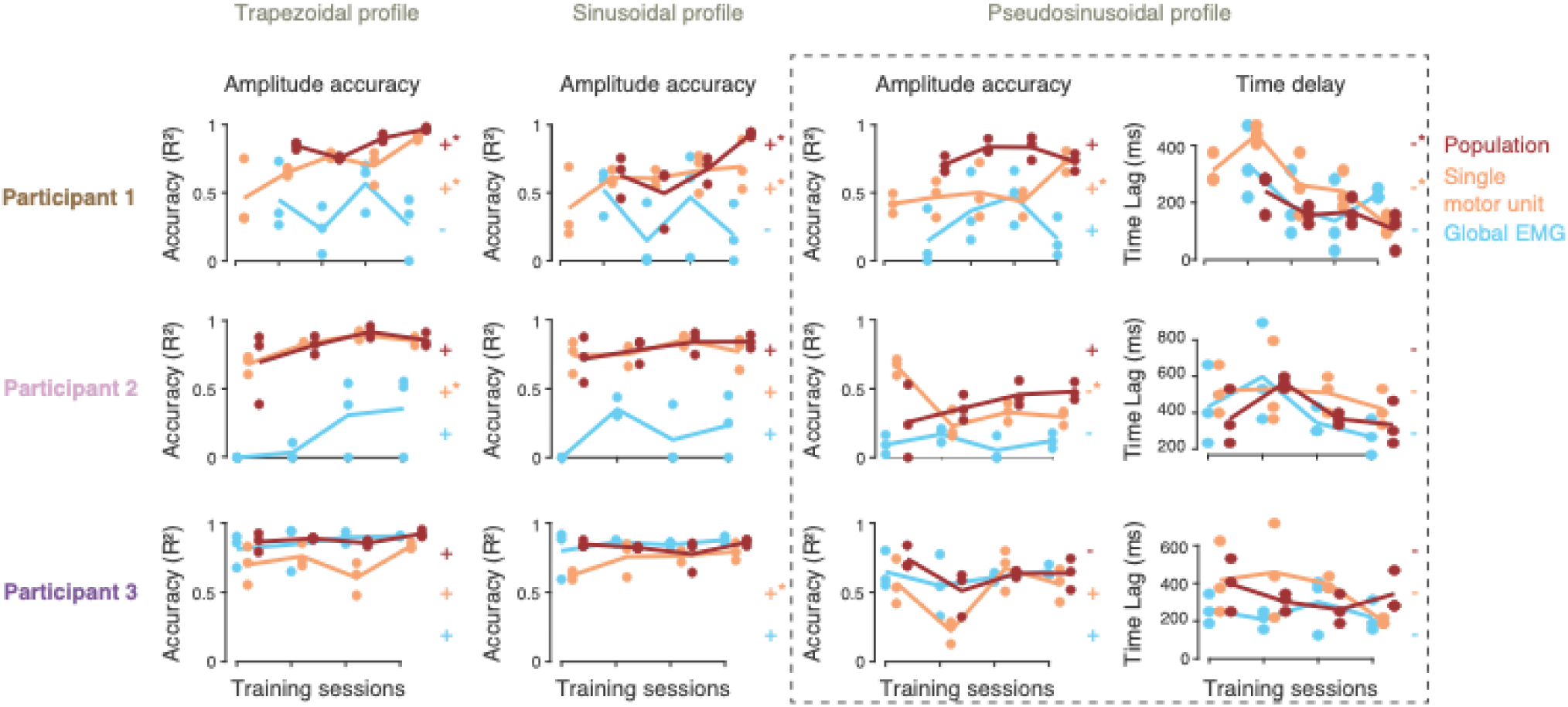
Tracking performance across practice sessions spanning four to twelve weeks for all three participants, including compensation for control delays. Like Figure 3, but for the pseudosinusoidal profile we also calculated a lag-compensated *R*^2^(left column, “Amplitude accuracy”) by dividing the full profile into semi-sinusoidal segments and then calculating the tracking accuracy after aligning the online profile to the segment using cross-correlation; right, mean estimated time delay across all segments (“Time delay”). We fitted linear regressors to the across-day performance trends; + indicates a positive slope, - a negative slope, no sign a zero slope, * a statistically significant change (*P*<0.05, *t*-test); signs are colour-matched with the control signals.

**Supplementary Figure 6.**
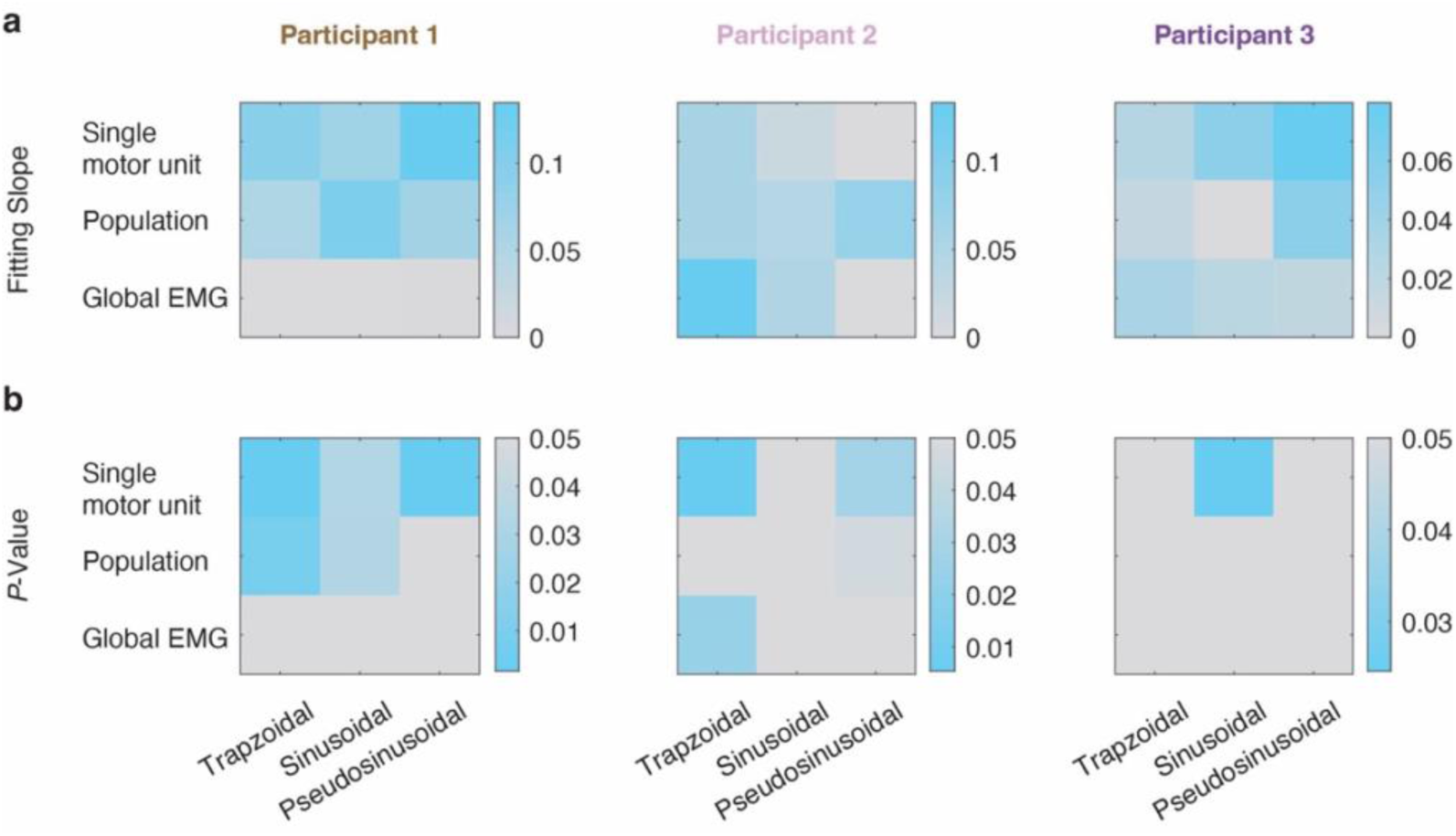
Effect of practice on one-dimensional motor unit control. **a**. Slope of the linear fit to the tracking accuracy as a function of days, for each control signal (row) and tracking profile (bottom). Blue, positive slopes; grey, negative or zero slopes. **b**. *P*-value of the linear fits. Blue, *P*<0.05; grey, *P*≥0.05.

**Supplementary Figure 7.**
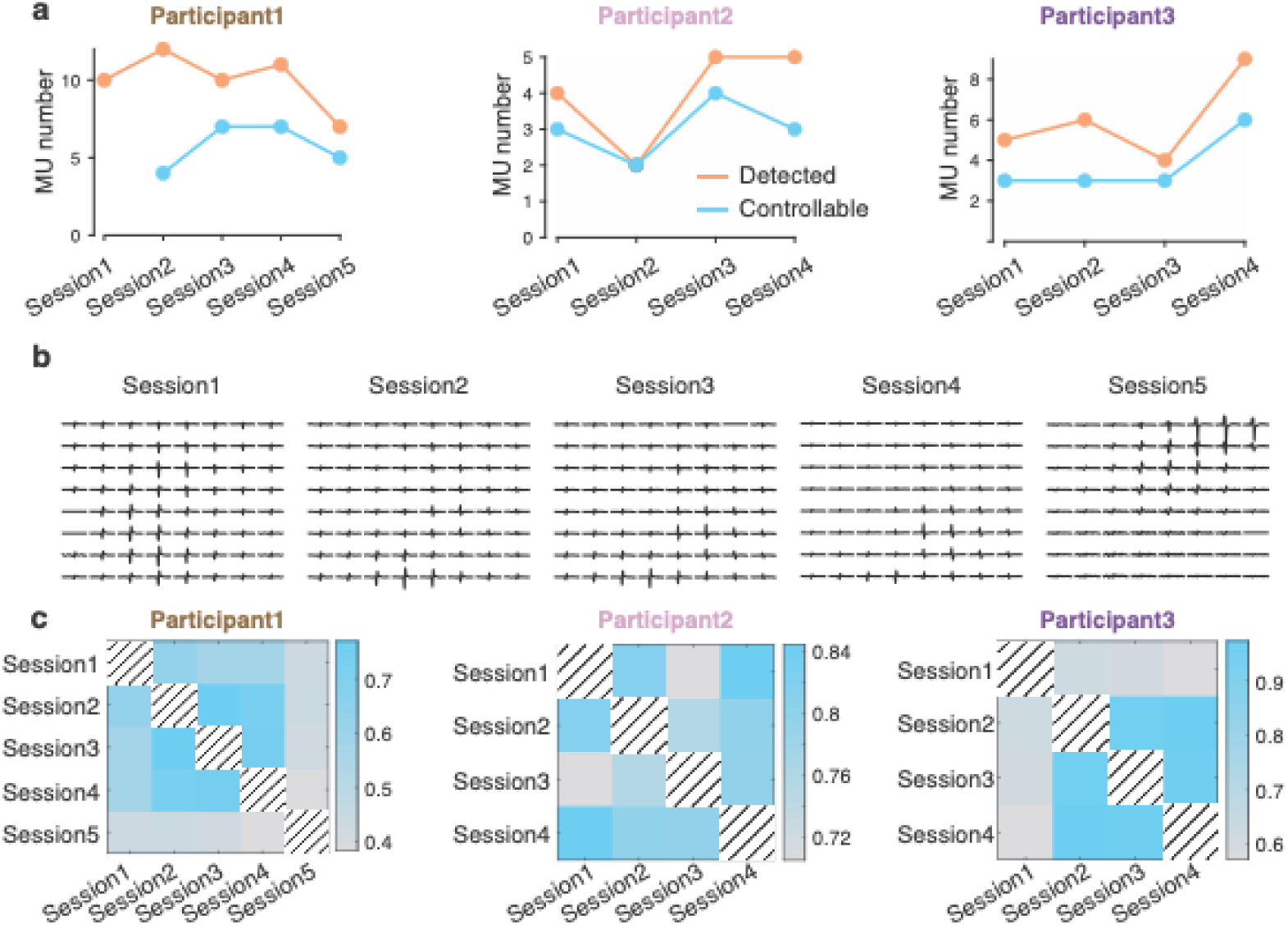
Motor unit characteristics across multiple sessions of practice. **a**. Total number of reliably decomposed motor units after postprocessing (orange), and number of those that were controllable online (blue), for each session and participant. Note: we did not assess online the number of controllable units on session 1 for P1. **b.** Motor unit action potential across all electrodes of one 8×8 HDsEMG array for the controlled motor unit across all sessions for P1. **c**. Pearson’s correlation coefficient between the motor unit action potentials of the controlled motor unit across sessions, indicating that participants did not always control the same unit.

**Supplementary Figure 8.**
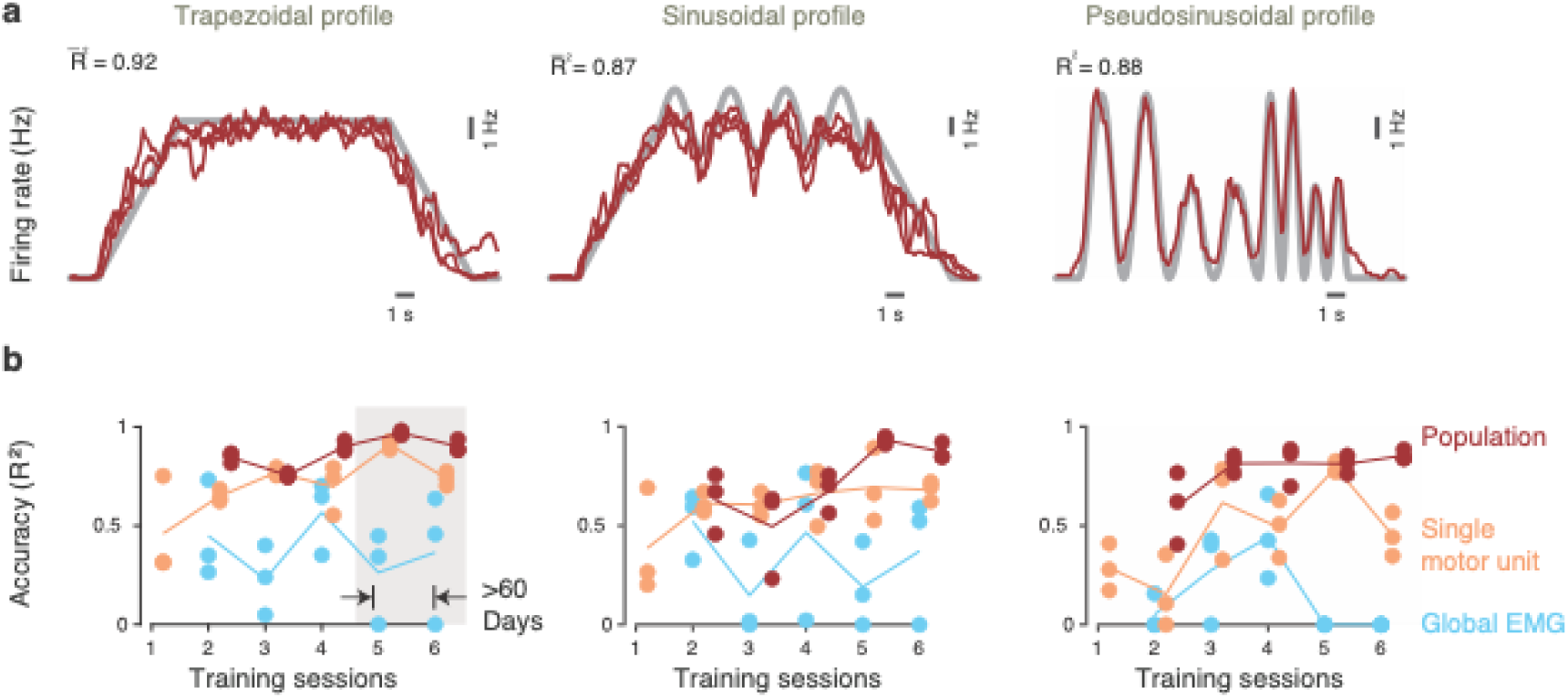
Retention of intuitive motor unit control more than 60 days after training. **a**. Online control during an additional sixth session from P1 as he tracked the three profiles using the activity of a population of motor units. Tracking accuracy was measured with the coefficient of determination (*R*^2^); *R^-^*^2^is the average tracking accuracy across multiple trials. **b**. Comparison of tracking performance during the “retention session” (session 6) with that during training (sessions 1 to 5).

**Supplementary Figure 9.**
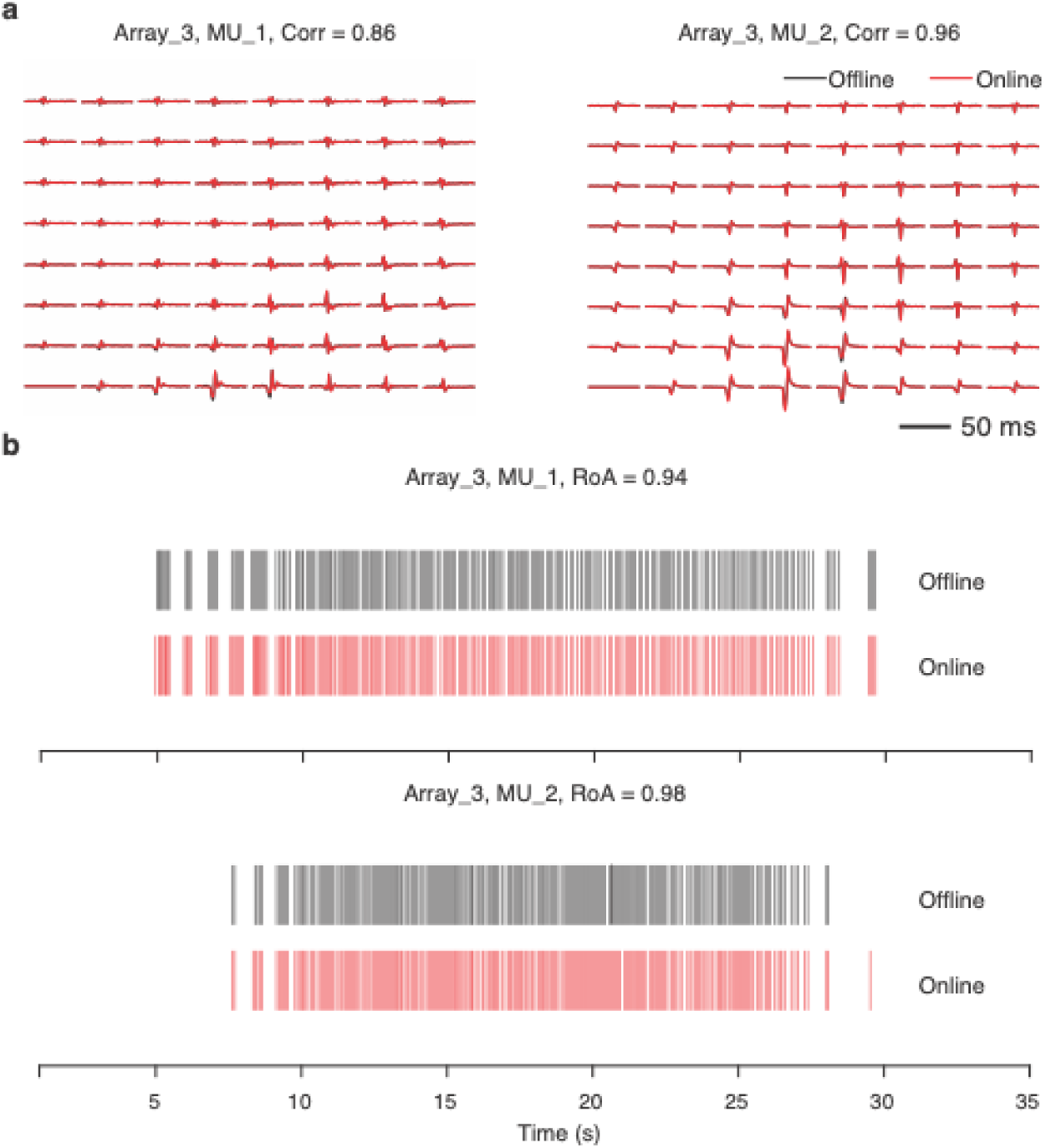
Offline validation of online motor unit decomposition. Even though the fact that participants could intuitively control the activity of single motor units and of population of motor units indicates that HDsEMG decomposition was accurate, we further validated the accuracy of our online detection of motor units by comparing it to offline decomposition. a. Example comparison of motor unit action potentials waveforms between offline and online decomposition for two motor units from P1 detected across one HDsEMG array. Black lines, motor unit action potentials identified during offline decomposition; red lines, same for online control; Corr, correlation across all channels b. Comparison of motor unit discharge trains during offline and online decomposition. RoA, rate of agreement; black lines, motor unit discharges identified through offline decomposition; red same for online decomposition.

## Supplementary videos

The following Supplementary videos can be accessed at Zenodo: https://doi.org/10.5281/zenodo.17841392

**Supplementary Video 1.** Example of intuitive control of a population of motor units.

**Supplementary Video 2.** Example of two-dimensional cursor navigation.

**Supplementary Video 3.** Example of three-dimensional motor unit control during the three bars task.

**Supplementary Video 4.** Example of playing Pong with one motor unit.

**Supplementary Video 5.** Example of playing Snake with two motor units.

## Footnotes

* Motor unit refers to a spinal motoneuron and the muscle fibres it innervates. Due to the reliability of the neuromuscular junction through which a motoneuron innervates the muscle fibres, each motoneuron action potential becomes a propagating motor unit action potential.

## References

1 Safdarian, M. et al. Global, regional, and national burden of spinal cord injury, 1990–2019: a systematic analysis for the Global Burden of Disease Study 2019. The Lancet Neurology 22, 1026–1047 (2023).

2 Anderson, K. D. Targeting recovery: priorities of the spinal cord-injured population. Journal of neurotrauma 21, 1371–1383 (2004).

3 Barra, B. et al. Epidural electrical stimulation of the cervical dorsal roots restores voluntary upper limb control in paralyzed monkeys. Nature Neuroscience 25, 924–934 (2022).

4 Badi, M. et al. Intrafascicular peripheral nerve stimulation produces fine functional hand movements in primates. Science Translational Medicine 13, eabg6463 (2021).

5 Ajiboye, A. B. et al. Restoration of reaching and grasping movements through brain-controlled muscle stimulation in a person with tetraplegia: a proof-of-concept demonstration. The Lancet 389, 1821–1830 (2017).

6 Bouton, C. E. et al. Restoring cortical control of functional movement in a human with quadriplegia. Nature 533, 247–250 (2016).

7 Ethier, C., Oby, E. R., Bauman, M. J. & Miller, L. E. Restoration of grasp following paralysis through brain-controlled stimulation of muscles. Nature 485, 368–371 (2012).

8 Hochberg, L. R. et al. Neuronal ensemble control of prosthetic devices by a human with tetraplegia. Nature 442, 164–171 (2006).

9 Collinger, J. L. et al. High-performance neuroprosthetic control by an individual with tetraplegia. The Lancet 381, 557–564 (2013).

10 Willett, F. R., Avansino, D. T., Hochberg, L. R., Henderson, J. M. & Shenoy, K. V. High-performance brain-to-text communication via handwriting. Nature 593, 249–254 (2021). 10.1038/s41586-021-03506-2

11 Dalrymple, A. N., Jones, S. T., Fallon, J. B., Shepherd, R. K. & Weber, D. J. Overcoming failure: improving acceptance and success of implanted neural interfaces. Bioelectronic Medicine 11, 6 (2025).

12 Balbinot, G. et al. Segmental motor recovery after cervical spinal cord injury relates to density and integrity of corticospinal tract projections. Nature communications 14, 723 (2023).

13 Oliveira, D. S. et al. A direct spinal cord–computer interface enables the control of the paralysed hand in spinal cord injury. Brain 147, 3583–3595 (2024).

14 Ting, J. E. et al. Sensing and decoding the neural drive to paralyzed muscles during attempted movements of a person with tetraplegia using a sleeve array. Journal of neurophysiology 126, 2104–2118 (2021).

15 Sîmpetru, R. C. et al. MyoGestic: EMG interfacing framework for decoding multiple spared motor dimensions in individuals with neural lesions. Science Advances 11, eads9150 (2025).

16 Lu, Z., Stampas, A., Francisco, G. E. & Zhou, P. Offline and online myoelectric pattern recognition analysis and real-time control of a robotic hand after spinal cord injury. Journal of neural engineering 16, 036018 (2019).

17 Yun, Y. et al. in 2017 IEEE international conference on robotics and automation (ICRA). 2904–2910 (IEEE).

18 Balbinot, G. et al. Properties of the surface electromyogram following traumatic spinal cord injury: a scoping review. Journal of neuroengineering and rehabilitation 18, 105 (2021).

19 Sköld, C., Levi, R. & Seiger, Å. Spasticity after traumatic spinal cord injury: nature, severity, and location. Archives of physical medicine and rehabilitation 80, 1548–1557 (1999).

20 Maynard, F., Karunas, R. & Waring 3rd, W. Epidemiology of spasticity following traumatic spinal cord injury. Archives of physical medicine and rehabilitation 71, 566–569 (1990).

21 Gorassini, M. A., Knash, M. E., Harvey, P. J., Bennett, D. J. & Yang, J. F. Role of motoneurons in the generation of muscle spasms after spinal cord injury. Brain 127, 2247–2258 (2004).

22 Zijdewind, I. & Thomas, C. K. Spontaneous motor unit behavior in human thenar muscles after spinal cord injury. Muscle & Nerve: Official Journal of the American Association of Electrodiagnostic Medicine 24, 952–962 (2001).

23 de Oliveira, D. S. et al. Spared Motor Neurons Enable the Control of a Robotic Sixth-Finger for Assistive Grasping in Tetraplegia. medRxiv, 2025.2002. 2007.25321673 (2025).

24 Yang, X. et al. Non-Invasive Neural Interfacing for Tetraplegic Individuals Using Residual Motor Neuron Activity Decoded At the Forearm or Wrist. IEEE Journal of Biomedical and Health Informatics (2025).

25 Holobar, A. & Zazula, D. Multichannel blind source separation using convolution kernel compensation. IEEE Transactions on Signal Processing 55, 4487–4496 (2007).

26 Negro, F., Muceli, S., Castronovo, A. M., Holobar, A. & Farina, D. Multi-channel intramuscular and surface EMG decomposition by convolutive blind source separation. J Neural Eng 13, 026027 (2016). 10.1088/1741-2560/13/2/026027

27 Farina, D. & Negro, F. Common synaptic input to motor neurons, motor unit synchronization, and force control. Exercise and sport sciences reviews 43, 23–33 (2015).

28 Farina, D., Negro, F. & Dideriksen, J. L. The effective neural drive to muscles is the common synaptic input to motor neurons. The Journal of physiology 592, 3427–3441 (2014).

29. contributors, W. Pong, <https://en.wikipedia.org/w/index.php?title=Pong&oldid=1323327675> (2025).

30. contributors, W. Snake (1998 video game), <https://en.wikipedia.org/w/index.php?title=Snake_(1998_video_game)&oldid=1317837783> (2025).

31 Negro, F., Muceli, S., Castronovo, A. M., Holobar, A. & Farina, D. Multi-channel intramuscular and surface EMG decomposition by convolutive blind source separation. Journal of neural engineering 13, 026027 (2016).

32 Rossato, J. et al. I-Spin live, an open-source software based on blind-source separation for real-time decoding of motor unit activity in humans. Elife 12, RP88670 (2024).

33 Hutson, T. H. & Di Giovanni, S. The translational landscape in spinal cord injury: focus on neuroplasticity and regeneration. Nature Reviews Neurology 15, 732–745 (2019).

34 Lin, D. J., Cramer, S. C., Boyne, P., Khatri, P. & Krakauer, J. W. High-Dose, High-Intensity Stroke Rehabilitation: Why Aren’t We Giving It? Stroke 56, 1351–1364 (2025).

35 Dawson, A. et al. High-dose high-intensity arm neurorehabilitation in chronic stroke improves general motor control. medRxiv, 2025.2011. 2018.25340491 (2025).

36 Makin, T. R. & Krakauer, J. W. Against cortical reorganisation. elife 12, e84716 (2023).

37 Edelman, B. J. et al. Noninvasive neuroimaging enhances continuous neural tracking for robotic device control. Science robotics 4, eaaw6844 (2019).

38 Lee, J. Y. et al. Brain–computer interface control with artificial intelligence copilots. Nature Machine Intelligence, 1–14 (2025).

39 Hwang, H.-J., Kwon, K. & Im, C.-H. Neurofeedback-based motor imagery training for brain–computer interface (BCI). Journal of neuroscience methods 179, 150–156 (2009).

40 Ghavampour, A. et al. A paradigm to study the learning of muscle activity patterns outside of the natural repertoire. Journal of Neurophysiology 134, 347–360 (2025).

41 Sadtler, P. T. et al. Neural constraints on learning. Nature 512, 423–426 (2014).

42 Willsey, M. S. et al. A high-performance brain–computer interface for finger decoding and quadcopter game control in an individual with paralysis. Nature Medicine 31, 96–104 (2025).

43 Carmena, J. M. et al. Learning to control a brain–machine interface for reaching and grasping by primates. PLoS biology 1, e42 (2003).

44 Li, D., Chen, C., Zhu, K., Guo, R. & Shull, P. B. Integrating Motor Unit Activity With Deep Learning for Real-Time, Simultaneous and Proportional Wrist Angle and Grasp Force Estimation. IEEE Transactions on Biomedical Engineering (2025).

45 Schone, H. R. et al. Biomimetic versus arbitrary motor control strategies for bionic hand skill learning. Nature human behaviour 8, 1108–1123 (2024).

46 Farina, D., Holobar, A., Merletti, R. & Enoka, R. M. Decoding the neural drive to muscles from the surface electromyogram. Clinical neurophysiology 121, 1616–1623 (2010).

47. Day, S. Important factors in surface EMG measurement. Bortec Biomedical Ltd publishers, 1–17 (2002).

48 Farina, D., Mesin, L., Martina, S. & Merletti, R. A surface EMG generation model with multilayer cylindrical description of the volume conductor. IEEE Transactions on Biomedical Engineering 51, 415–426 (2004).

49 Braun, D. I. et al. NeurOne: High-performance motor unit-computer interface for the paralyzed. medRxiv, 2023.2009. 2025.23295902 (2023).

50 Fuglevand, A. J., Lester, R. A. & Johns, R. K. Distinguishing intrinsic from extrinsic factors underlying firing rate saturation in human motor units. Journal of neurophysiology 113, 1310–1322 (2015).

51 Hug, F., Avrillon, S., Ibáñez, J. & Farina, D. Common synaptic input, synergies and size principle: Control of spinal motor neurons for movement generation. The Journal of physiology 601, 11–20 (2023).

52 Del Vecchio, A. et al. Tutorial: Analysis of motor unit discharge characteristics from high-density surface EMG signals. Journal of Electromyography and Kinesiology 53, 102426 (2020).

53 Barsakcioglu, D. Y., Bräcklein, M., Holobar, A. & Farina, D. Control of spinal motoneurons by feedback from a non-invasive real-time interface. IEEE Transactions on Biomedical Engineering 68, 926–935 (2020).

54 Clancy, E. A. & Hogan, N. Probability density of the surface electromyogram and its relation to amplitude detectors. IEEE Transactions on Biomedical Engineering 46, 730–739 (2002).

55 Montgomery, D. C., Peck, E. A. & Vining, G. G. Introduction to linear regression analysis. (John Wiley & Sons, 2021).

